# Cardiometabolic risk and structural brain development in a large community-based U.S. cohort

**DOI:** 10.1101/2025.10.23.25338683

**Authors:** Dani Beck, Lars T. Westlye, Christian K. Tamnes

**Author notes:** Corresponding author: Dani Beck. Co-author.

## Abstract

**Objective:** Cardiometabolic risk factors are already detectable in childhood and adolescence, but their relation to the developing brain remains unclear. The current study tested whether poorer cardiometabolic health is associated with brain structure and microstructure development in 10–17-year-old youth.

**Methods:** Using the Adolescent Brain Cognitive Development Study, we analysed data from 3,527 participants with 4,433 observations across three waves (single wave: n = 2,745; two waves: n = 658 participants; three waves: n = 124 participants). We related anthropometric (body-mass index, waist circumference), cardiovascular (systolic and diastolic blood pressure, resting heart rate), and metabolic (haemoglobin A1c, high-density lipoprotein cholesterol) indices to global cortical thickness and surface area, and to white matter fractional anisotropy and mean diffusivity. Bayesian multilevel models were fitted to estimate main and time-interaction effects, and sensitivity analyses tested within-person change, prospective prediction to the next wave, and replaced chronological age with puberty status.

**Results:** Higher resting heart rate was associated with higher mean diffusivity, an association that strengthened over time. Higher waist circumference was associated with larger surface area. Other cardiometabolic measures favoured the null, and sensitivity analyses provided little evidence that wave-to-wave changes in cardiometabolic health tracked contemporaneous brain change or predicted subsequent brain structure.

**Conclusion:** Across late childhood and adolescence, brain architecture appears largely insensitive to short-term variation in cardiometabolic risk indices.

## 1. Introduction

Cardiometabolic risk factors (CMRs) and brain structural integrity are strongly connected. In adults, higher adiposity^1,2^, dyslipidemia^3^, impaired glucose regulation^3^, and elevated blood pressure^3^ relate to macro- and micro-structural brain metrics and higher brain age, suggesting shared common biological pathways. Yet, the antecedents of these relations and the risk they pose begin much earlier in life. Cardiometabolic risk factors are already detectable during childhood and adolescence, with rising rates of hypertension^4,5^ and obesity^6,7^ linked to compromised brain structure. Guided by calls for larger samples and longitudinal designs to better characterise cardiometabolic risk in youth^8^, the current study aims to test whether poor cardiometabolic profiles of US children aged 10-17 years old is associated with poorer brain structural integrity across three waves of data.

Cardiometabolic health in U.S. youth has significantly worsened in the last two decades^9^. Approximately 19% of U.S. children meet criteria for obesity^10^, and rates of elevated blood pressure and hypertension have also increased^4,6,11,12^. From 2013-2016, 7.1% of youth aged 8–17 years had elevated blood pressure and 3.5% met criteria for hypertension^13,14^. These figures continue to rise and may have cumulative effects on the developing brain.

Cross-sectional magnetic resonance imaging (MRI) studies have consistently reported associations between childhood obesity and differences in cortical thickness^7^, total grey matter volume^15–19^, and white matter architecture^20,21^, and although longitudinal studies report comparable findings, such evidence is limited^22,23^. The directionality of adiposity-related effects is relatively consistent for cortical thickness – with higher BMI generally associated with thinner cortex^7,19^ – though findings for grey matter volume and white matter microstructure are more mixed. Evidence linking blood pressure (BP) to brain structure in youth is comparatively limited. Cross-sectional analyses found higher diastolic BP associations with smaller total and grey matter volume, but weaker or null associations for systolic BP and white matter microstructure (WMM)^24^. Neural correlates of dyslipidemia and glycaemic regulation are even less well characterised. Although high-density lipoprotein (HDL) cholesterol and haemoglobin A1c (HbA1c) are routinely assessed in paediatric care and tracked into adulthood, few studies have linked them to brain outcomes. Some have reported null associations^21^, while others suggest that elevated lipid levels or poorer overall cardiometabolic health cross-sectionally relate to smaller cortical volumes^25^. Critical gaps remain, with most prior work being cross-sectional, based on modest samples (Ns ranging from approximately 30 to 400 in many studies, though some larger cohorts exist^19,21,22^, conducted predominantly in children and adolescents aged 8–18 years), and with very few examining lipid- or glycemia-specific brain associations in youth.

The current study aims to fill gaps in the literature by leveraging the large, longitudinal ABCD Study to test whether poorer cardiometabolic health in U.S. youth is associated with alterations in brain structure and WMM across three waves of data collection (the two-, four-, and six-year follow-up assessments, each separated by approximately two years, covering ages 10.6–17.8 years with mean ages of 11.97, 14.26, and 16.08 years at each wave respectively). We examine anthropometric (BMI, waist circumference (WC)), cardiovascular (systolic and diastolic BP, resting heart rate), and metabolic (HbA1c, HDL cholesterol) indices in relation to cortical thickness (CT), surface area (SA), and diffusion-derived metrics – fractional anisotropy and mean diffusivity (MD).

BMI and waist circumference are indices of overall and central adiposity respectively, with elevated values associated with increased cardiometabolic and metabolic disease risk^11,26^. Systolic and diastolic blood pressure reflect the force exerted on arterial walls during and between heartbeats, with elevated levels indicating hypertension risk even in youth^12^. Resting heart rate is a marker of autonomic nervous system function and cardiorespiratory fitness, with higher values associated with poorer cardiovascular health^27^. HbA1c reflects average blood glucose levels over approximately three months and is used to screen for prediabetes and diabetes^28^, while HDL cholesterol is a marker of lipid metabolism, with lower levels associated with increased cardiovascular risk^29^.

We fit Bayesian multilevel models to estimate associations while accounting for sex, age, and within-person repeated measures. Our primary analysis tests whether cardiometabolic health in late childhood-to-late adolescence are associated with brain structure at the same visit and over time. As sensitivity analyses, we (i) use a change-score model (Δbrain ∼ Δcardiometabolic) to isolate within-individual covariation across waves, (ii) use a lagged prospective model (brain_t+1_ ∼ cardiometabolic_t_ + brain_t_) to test whether cardiometabolic status predicts subsequent brain change beyond prior brain level, and (iii) replace chronological age with pubertal development given its strong correlation with age and its closer proximity to biological maturation.

We hypothesise that higher cardiometabolic risk – reflected in higher BMI, WC, systolic and diastolic BP, heart rate, and HbA1c, and lower HDL cholesterol – will relate to globally thinner cortex and poorer WMM (lower FA, higher MD) at the same visit, with minimal or null effects for surface area given its early developmental determination. We further expect these associations to strengthen across waves based on accumulated risk and few existing longitudinal studies. In change-score models, wave-to-wave increases in cardiometabolic risk are expected to track contemporaneous cortical and white matter change, while in lagged models, higher cardiometabolic risk at one wave is anticipated to predict differences in brain structure at the next wave – specifically thinner cortex, lower FA, and higher MD – beyond prior brain level. We expect that replacing chronological age with pubertal development status will clarify whether observed associations are driven by biological maturation rather than chronological age per se, and whether any cardiometabolic effects persist independently of maturational stage.

## 2. Methods

### 2.1. Sample description and ethical approval

The ABCD Study ®^30^ is an ongoing longitudinal study comprising ∼11,800 children and adolescents. Data used in the present study were drawn from the ABCD Study release 6.0, containing data from baseline up until the six-year visit (nbdc-datahub.org/abcd-release-6-0).

All ABCD Study data is stored in the NIH Brain Development Cohorts (NBDC) Data Hub, which is available for authorised users with approved Data Use Certification (DUC) (lead investigator: Westlye). The 6.0 release has been assigned the DOI <u>10.82525/jy7n-g441</u>. The Institutional Review Board (IRB) at the University of California, San Diego, approved all aspects of the ABCD Study^31^. Parents or guardians provided written consent, while the child provided written assent. The current study was conducted in line with the Declaration of Helsinki and was approved by the Norwegian Regional Committee for Medical and Health Research Ethics (REK 2019/943).

The initial sample included ∼11,800 participants. The ABCD Study exclusion criteria included non-English proficiency, general MRI contraindications, a history of a major neurological disorder, traumatic brain injury, extreme premature birth (<28 weeks gestational age), a diagnosis of schizophrenia, intellectual disability, moderate to severe autism spectrum disorder, or substance abuse disorder, and participants not meeting these criteria were included in the current study (see Karcher et al. (2018)^32^ for more detail).

The analytic sample was substantially reduced due to limited availability of CMR measures and the requirement that CMR and brain MRI data be available at the same assessment waves. Of the initial cohort, ∼4,000 participants with ∼5,000 observations had overlapping, quality-controlled CMR and MRI data at the two-, four-, or six-year follow-up sessions. Analyses were restricted to these matched sessions. Standard ABCD quality control procedures were applied to MRI data, and additional quality assurance and plausibility checks were conducted for all CMR measures. One unrelated participant per family was retained to minimise confounding due to familial relatedness. Full details of step-by-step sample selection and quality assurance procedures are provided in Supplementary Information (SI); SI Section 1.

The final sample consisted of 3527 unique participants with 4433 observations (52.8% male) at mean age 14.6 years (SD = 1.73, range 10.6–17.8), with mixed cross-sectional and longitudinal CMR and brain MRI data from the two- (N = 970), four- (N = 1,493), and six-year (N = 1,970) follow-up time points. The 4433 observations include 2745 single observation data points from any of the three waves, 1316 observations (N = 658) representing longitudinal data from any two waves, and 372 observations (N = 124) from participants with data in each of the three waves. Demographic information pertaining to the analytic sample and across participation groups can be found in Table 1 and illustrated in Figure 1 below. Demographic and health characteristics of the analytic sample compared to the full ABCD cohort is provided in SI Table 1. Formal attrition analyses comparing participation groups are reported in SI Figure 1. Intraclass correlation coefficients and mean within-person standard deviations for each cardiometabolic measure are reported in SI Table 2.

**Figure 1.**
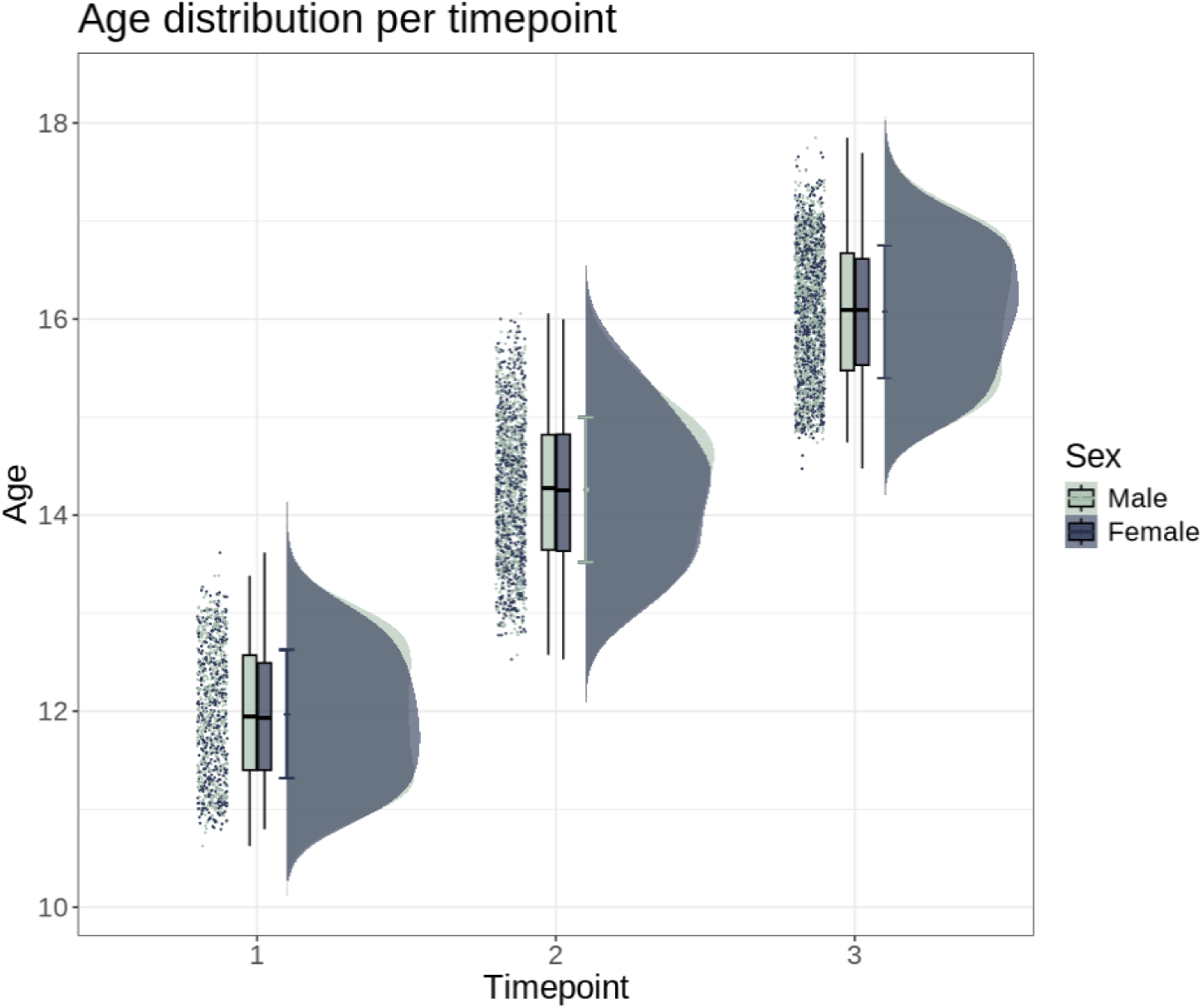
Sample age and sex distribution at timepoint 1, 2, and 3 (year two, four, and six) of the ABCD Study.

**Table 1.**
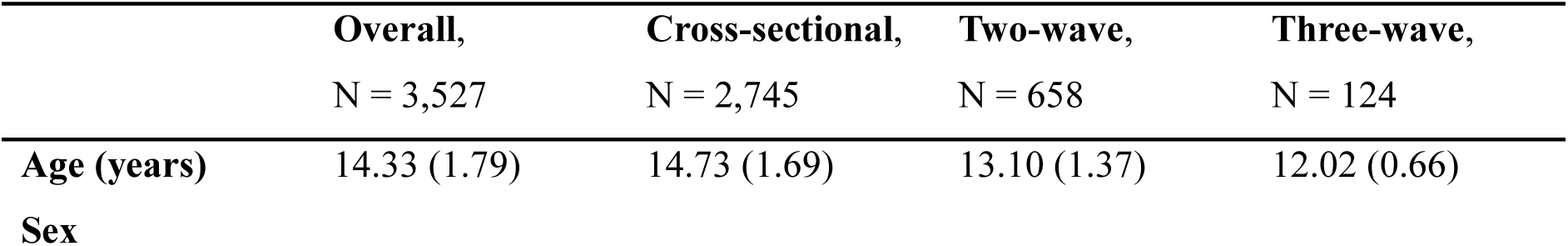
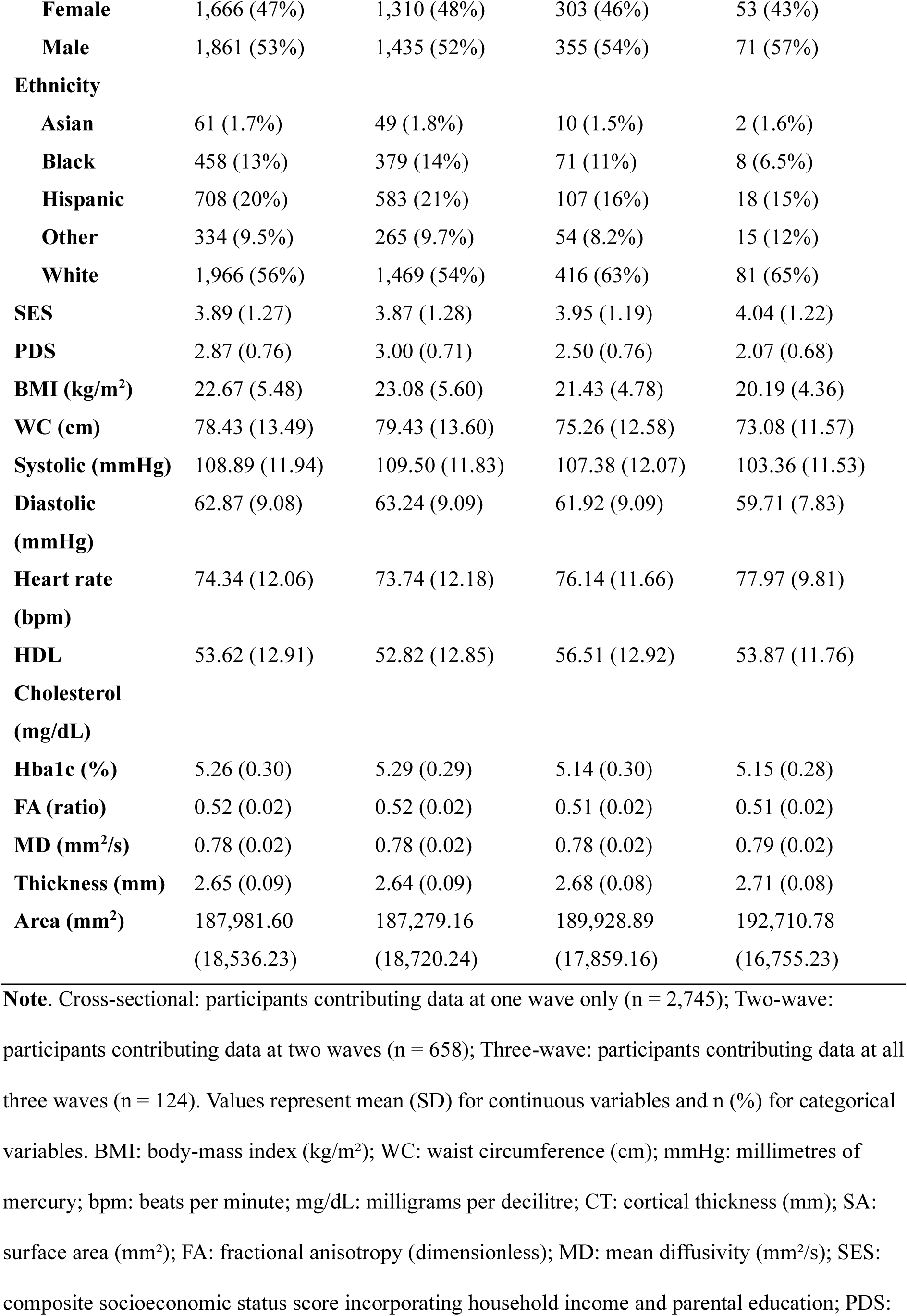
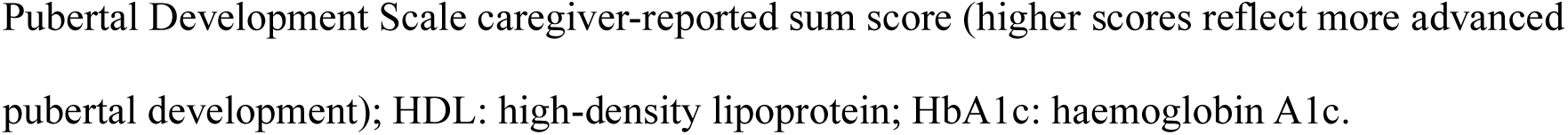
Demographic, cardiometabolic, and neuroimaging characteristics of the analytic sample stratified by participation pattern.

### 2.2. MRI acquisition and processing

Neuroimaging data were acquired at 21 different sites (using 34 scanners) and processed by the ABCD Study team. A 3-T Siemens Prisma, General Electric 750, or Phillips scanner was used for data acquisition. Protocols used for data acquisition and processing are described elsewhere^30,33^ and available in SI Section 2. Briefly, two-, four-, and six-year follow-up structural grey matter measures of global cortical thickness (CT) and surface area (SA), and diffusion tensor imaging measures of white matter fractional anisotropy (FA) and mean diffusivity (MD) were used for the current study. Cortical surface reconstruction and subcortical segmentation was performed with FreeSurfer v7.1.1^34,35^. White matter microstructural measures were generated using AtlasTrack, a probabilistic atlas-based method for automated segmentation of white matter fibre tracts^36^. Inner shell DTI measures (b ≤ 1000 s/mm²) were used for FA and MD, following Hagler et al. (2019)^37^, as these yield values most comparable to the broader DTI literature. Following procedures of quality assurance (see SI Section 1), harmonisation of multi-scanner effects was carried out using *longCombat*^38^ (see SI Section 3 and SI Figure 2). Brain trajectories for each brain measure across the three timepoints is illustrated in SI Figures 3 and 4.

### 2.3. Cardiometabolic risk factors

Anthropometric markers of height, weight, waist circumference (WC), and body-mass index (BMI) are collected during visitation. BMI is calculated as weight (kg) / (height (cm) / 100)², following conversion from inches to centimetres (inches × 2.54) for height and from pounds to kilograms (lbs / 2.2046) for weight. WC is measured in cm. Obesity is defined as a BMI at or above the 95th percentile for children and teens of the same age and sex^26^.

Prior to measurement of blood pressure, participants sat in a chair for 5 min in a quiet environment. The right arm was rested palm face up on a table, and feet were positioned flat on the floor, legs uncrossed. Blood pressure was calculated using the mean of three measurements separated by a 60s interval using a factory-calibrated, Omron blood pressure monitor (MicroLife USA, Inc.; Dunedin, FL). Hypertensive blood pressure was defined using age- and sex-appropriate percentiles from paediatric guidelines for elevated blood pressure^12^ (see SI Table 3). Heart rate (measured as beats per minute while resting) is also collected, with normal ranges of 70-100 for children 6–12 years and 60-100 for adolescents (13–18 years).

Haemoglobin A1c (HbA1c) was measured in venous blood as a measure of average blood sugar levels over the prior three months^39^ and is commonly used for screening and diagnosing of diabetes. According to the American Diabetes Association, participant tests are consistent with diabetes if they have HbA1c levels of 6.5% or more, and prediabetes in the 5.7-6.4% range^28,40^. Non-fasting High-Density Lipoprotein (HDL) cholesterol was measured in plasma, whereby low HDL cholesterol is defined as <40mg/dL^29^. Unlike triglycerides, which are substantially affected by recent food intake, HDL cholesterol concentrations are minimally influenced by fasting status, and non-fasting measurement is considered valid and widely used in large-scale paediatric and epidemiological research^41^.

A figure showing percentage of missing data for each CMR variable can be found in SI Figure 5. Density plots for each CMR overlayed by each wave and a spaghetti plot showing within-person trajectories of cardiometabolic risk measures across study waves are available in SI Figures 6 and 7, respectively. Intraclass correlation coefficients and mean within-person standard deviations for each CMR, including odds ratios from two logistic regression models predicting participation patterns are available in SI Figure 1 and SI Table 2. Quality assurance for each CMR is outlined SI Section 1 and visualised in SI Figures 8 (before outlier removal) and 9 (after outlier removal).

### 2.4. Statistical analysis

All analyses were carried out using R version 4.3.2^42^. To investigate the association (main effect) between each cardiometabolic measure and global metrics of CT, SA, FA, and MD, and to determine whether these associations vary across time points (interaction effect), we employed Bayesian multilevel models using the *brms*^43,44^ R-package, which is conceptually analogous to frequentist linear mixed effects models but operates within a Bayesian framework. Posterior distributions represent the full probability distribution of a parameter given the observed data and prior information, providing a complete characterisation of uncertainty rather than a single point estimate with a p-value. This approach was chosen because it allows principled quantification of evidence both in favour of and against an effect via Bayes Factors, provides full posterior distributions that characterise parameter uncertainty beyond point estimates, and is particularly well-suited to studies where null or small associations are plausible a priori^45^.

In each model, the brain metric was treated as the dependent variable; each CMR measure (systolic BP, diastolic BP, heart rate, WC, BMI, HbA1c, and HDL cholesterol) and its interaction with time point (TP; two-year, four-year, six-year session) were specified as fixed effects, age centred within timepoint (to reduce collinearity), sex, and socioeconomic status (SES) were included as covariates, and subject ID served as a random effect.

To regularise parameter estimates and mitigate false positives, we assigned a moderately weak prior centred on zero with a standard deviation of 1 (normal(0,1)) for all regression coefficients, reflecting a prior expectation of small effects but allowing sufficient flexibility for estimation. We ran each model for 10,000 iterations (5,000 warm-up, 5,000 post-warm-up) across four chains, following recommended practice for brms/Stan models to ensure convergence and thorough posterior sampling^43,44^. We set adapt delta = 0.99 to improve Hamiltonian Monte Carlo (HMC) stability and reduce divergent transitions, particularly important in models with random effects and modest collinearity.

In sensitivity analyses, we (i) fit a change-score specification on adjacent waves (Δbrain ∼ 0 + ΔCMR + ΔAge + Sex + SES + (1|ID)) to isolate within-person covariation while differencing out time-invariant confounding. We (ii) fit a lagged prospective model of the form brain _t+1_ ∼ CMR _t_ + brain _t_ + Age + Sex + SES + (1|ID) to test whether cardiometabolic status predicts subsequent brain beyond prior brain level. Finally, we (iii) replaced age with pubertal development status by means of pubertal development scale (PDS)^46^ caregiver-reported sum score across five items rated 1–4, where higher scores reflect more advanced pubertal development, used in previous research^47^. The PDS is a questionnaire designed to mimic the traditional Tanner staging assessment without the use of reference pictures. Due to previous research showing that youth tend to over-report their perceived physical development at younger ages^48^, the current study utilised caregiver PDS report.

While all primary models were fit using the full analytic dataset (3,527 participants), the change-score outcomes and lagged predictors are only defined for – and therefore informed by – the subset of participants with repeated measurements. In the present sample, 782 participants contributed longitudinal data, including 658 with data from two waves and 124 with data from all three waves; these participants therefore drive inference on within-person change and lagged effects.

For each coefficient of interest, we report the posterior mean (*β*) and 95% credible interval. In addition, we computed Bayes Factors (BF_12_) using the Savage-Dickey method^45^, which provide evidence ratios indicating how strongly the data support one hypothesis (null/M1 versus alternative/M2). We classify BFs into descriptive categories (e.g., extreme, very strong, strong, moderate, anecdotal, or none) following the guidelines in Jeffreys (1961)^49^ (see SI Table 4). The full results for all our models, including posterior estimates, credible intervals, and BFs, are provided in SI Tables 5-8.

## 3. Results

### 3.1. Cross-sectional and time-varying associations

Our primary model tested whether CMR levels are associated with concurrent brain structure, and whether these associations change across waves. Figure 2 shows the posterior distributions for associations between each CMR and global brain features.

**Figure 2.**
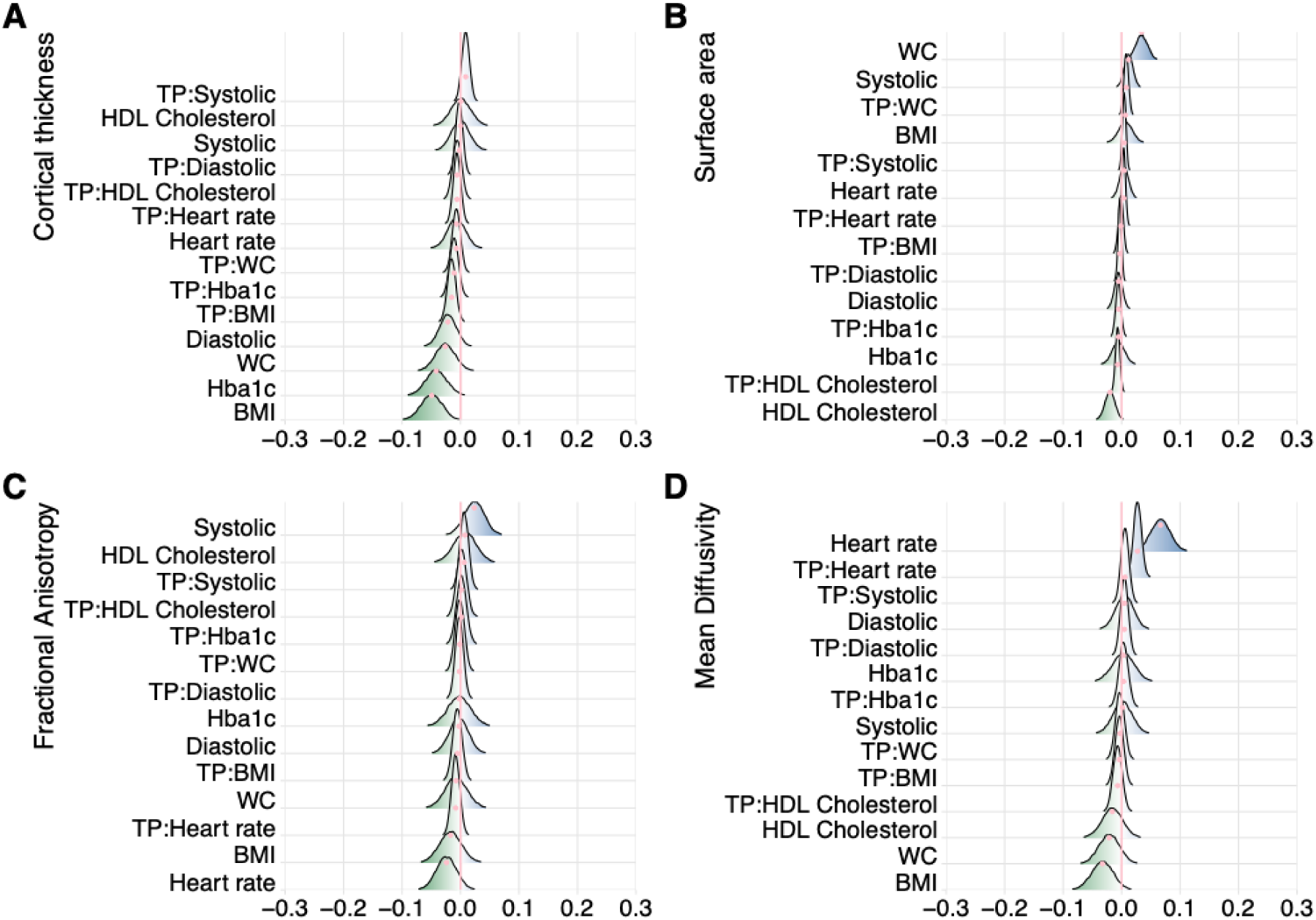
Associations between CMRs and global brain structure. The figure shows posterior distributions of the estimates of the standardised coefficient. Estimates for each CMR feature on cortical thickness (panel A) and surface area (panel B), fractional anisotropy (panel C), and mean diffusivity (panel D). Each row represents estimates from a separate model containing a single CMR predictor (main effect) or a single CMR × timepoint interaction term, alongside timepoint, age centred within timepoint, sex, SES, and subject ID as a random effect. Colour scale follows the directionality of evidence, with positive (blue) values indicating evidence in favour of positive associations (i.e., higher CMR and higher brain measure) and negative (green) values indicating evidence in favour of negative associations (i.e., higher CMR and lower brain measure). The width of the distribution represents the uncertainty of the parameter estimates. Systolic BP: systolic blood pressure; Diastolic BP: diastolic blood pressure; Heart rate: heart rate; HDL cholesterol: high-density lipoprotein cholesterol; Haemoglobin A1c: HbA1c; BMI: body-mass index; WC: waist circumference; TP: timepoint.

#### 3.1.1. Cortical thickness

Most main effects favoured the null: systolic BP (β = 0.000, [−0.030, 0.031], BF₁₂ = 65.19, strong for M1), diastolic BP (β = −0.022, [−0.049, 0.007], BF₁₂ = 22.96, strong for M1), heart rate (β = −0.007, [−0.037, 0.023], BF₁₂ = 56.61, strong for M1), HDL cholesterol (β = 0.000, [−0.033, 0.034], BF₁₂ = 58.91, strong for M1), HbA1c (β = −0.041, [−0.078, −0.007], BF₁₂ = 4.38, moderate for M1), and waist circumference (β = −0.026, [−0.059, 0.008], BF₁₂ = 19.01, strong for M1). BMI showed anecdotal evidence for a negative association with cortical thickness (β = −0.049, [−0.084, −0.015], BF₁₂ = 0.996). TP-interaction terms also favoured the null throughout (e.g., TP:diastolic BF₁₂ = 146.28, TP:heart rate BF₁₂ = 99.77, TP:HDL BF₁₂ = 104.88; all very strong–extreme for M1). The BMI×TP interaction was negative (β = −0.015 per wave, [−0.029, −0.001], BF₁₂ = 15.64, strong for M1).

#### 3.1.2. Surface area

Waist circumference showed moderate evidence for a positive association with surface area (β = 0.034, [0.015, 0.054], BF₁₂ = 0.265). All remaining main effects favoured the null, including HDL cholesterol (β = −0.020, [−0.037, −0.003], BF₁₂ = 8.062, moderate for M1), systolic BP (β = 0.012, [−0.003, 0.026], BF₁₂ = 38.43), diastolic BP (BF₁₂ = 110.37), heart rate (BF₁₂ = 111.34), BMI (BF₁₂ = 72.12), and HbA1c (BF₁₂ = 80.25). Interaction BFs were similarly large throughout.

#### 3.1.3. Fractional anisotropy

All estimates were small with strong or greater evidence for the null (e.g., systolic BF₁₂ = 21.70, BMI BF₁₂ = 33.84, heart rate BF₁₂ = 22.63). Time interactions also favoured M1 (e.g., TP:diastolic BF₁₂ = 135.92, TP:BMI BF₁₂ = 101.96).

#### 3.1.4. Mean diffusivity

Resting heart rate showed a positive association with MD (β = 0.066, [0.035, 0.099], BF₁₂ = 0.045, extreme for M2). The heart rate × timepoint interaction was also positive (β = 0.027 per wave, [0.014, 0.040], BF₁₂ = 0.068, strong for M2), indicating stronger associations at later timepoints. Other predictors largely favoured the null (e.g., systolic BP BF₁₂ = 61.50, diastolic BP BF₁₂ = 62.45, HbA1c BF₁₂ = 50.86, HDL cholesterol BF₁₂ = 39.36). WC showed strong evidence for the null (β = −0.021, [−0.056, 0.013], BF₁₂ = 27.72). BMI showed strong evidence for the null (β = −0.033, [−0.070, 0.002], BF₁₂ = 11.84).

### 3.2. Within-person co-variation

Our change score model tested whether wave-to-wave changes in cardiometabolic indices track contemporaneous changes in global brain metrics. Figure 3 shows the posterior distributions for associations between each ΔCMR and Δglobal brain features.

**Figure 3.**
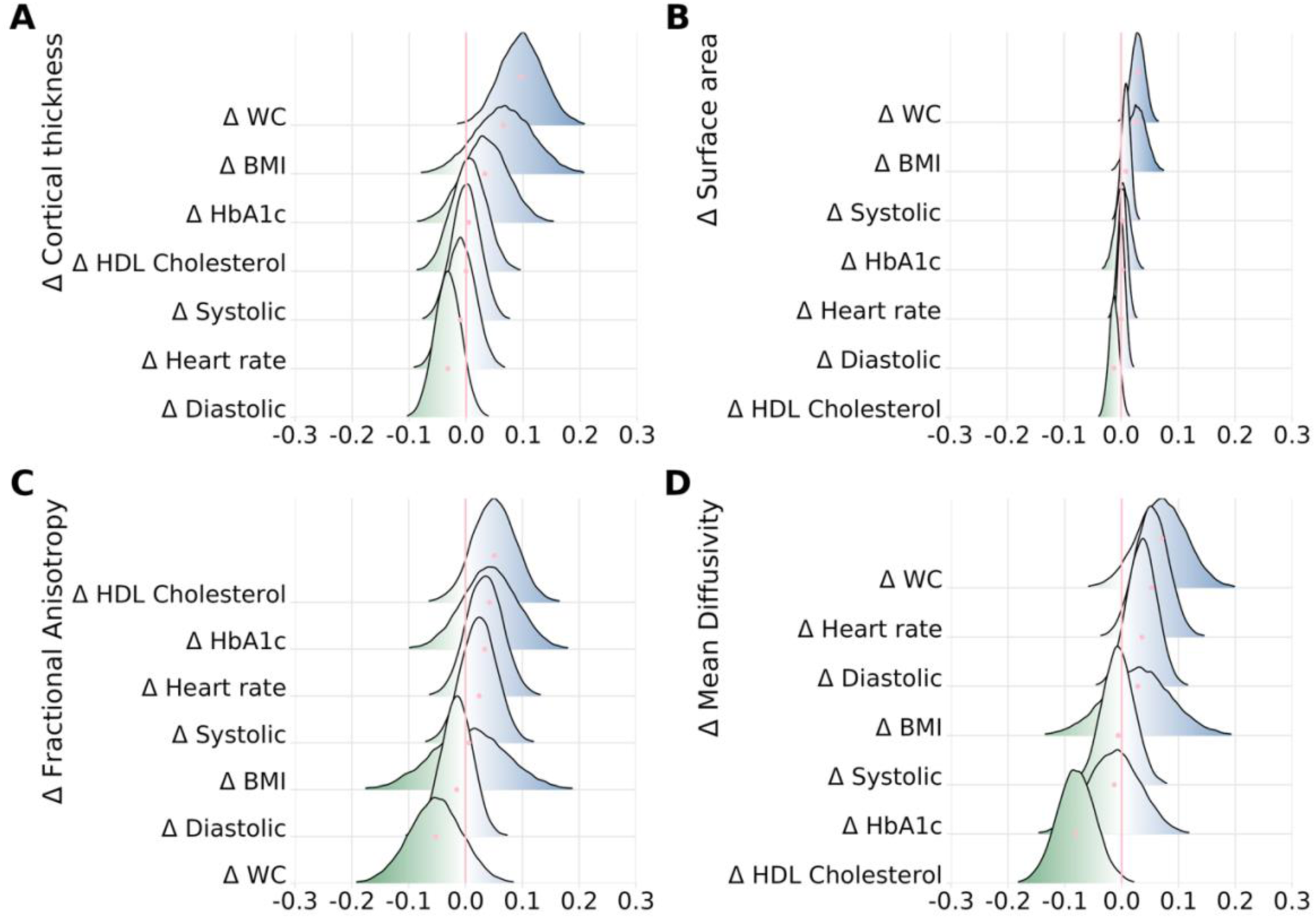
Within-person change-score associations between CMRs and global brain structure. Posterior distributions of the standardised coefficients from the change-score model Δbrain ∼ 0 + ΔCMR + ΔAge + Sex + SES + (1|ID), testing whether wave-to-wave changes in cardiometabolic risk track contemporaneous changes in (A) cortical thickness, (B) surface area, (C) fractional anisotropy, and (D) mean diffusivity. Colour indicates direction (blue = positive; green = negative); distribution width reflects uncertainty. Coefficients are interpreted as within-child effects after differencing out time-invariant confounds. Abbreviations as in Figure 2.

#### 3.2.1. Cortical thickness

Across CMRs, evidence generally favoured the null. Systolic BP (β = −0.000, [−0.049, 0.049], BF₁₂ = 39.73), heart rate (β = −0.010, [−0.062, 0.040], BF₁₂ = 34.93), and HDL cholesterol (β = 0.005, [−0.055, 0.063], BF₁₂ = 32.64) showed very strong evidence for the null. Diastolic BP (β = −0.032, [−0.077, 0.014], BF₁₂ = 16.86) and HbA1c (β = 0.033, [−0.044, 0.115], BF₁₂ = 18.21) showed strong evidence for the null. WC showed anecdotal evidence for the null (β = 0.097, [0.023, 0.174], BF₁₂ = 1.047).

#### 3.2.2. Surface area

Findings consistently favoured the null. Diastolic BP (β = 0.000, [−0.014, 0.015], BF₁₂ = 134.26) and heart rate (β = 0.003, [−0.014, 0.018], BF₁₂ = 117.09) showed extreme evidence for the null. Systolic BP (β = 0.009, [−0.007, 0.024], BF₁₂ = 71.17), HbA1c (β = 0.003, [−0.021, 0.028], BF₁₂ = 76.36), and HDL cholesterol (β = −0.012, [−0.030, 0.005], BF₁₂ = 43.52) showed very strong evidence for the null. BMI showed strong evidence for the null (β = 0.028, [−0.006, 0.059], BF₁₂ = 14.54). WC showed moderate evidence for the null (β = 0.030, [0.007, 0.054], BF₁₂ = 4.33).

#### 3.2.3. Fractional anisotropy

All change terms were small with strong to very strong evidence for no within-person association: systolic BP (β = 0.024, [−0.037, 0.084], BF₁₂ = 24.58), diastolic BP (β = −0.016, [−0.071, 0.039], BF₁₂ = 31.31), heart rate (β = 0.034, [−0.030, 0.096], BF₁₂ = 19.03), WC (β = −0.053, [−0.144, 0.040], BF₁₂ = 11.21), BMI (β = 0.006, [−0.124, 0.130], BF₁₂ = 14.90), HbA1c (β = 0.042, [−0.053, 0.135], BF₁₂ = 13.93), and HDL cholesterol (β = 0.051, [−0.022, 0.127], BF₁₂ = 10.65).

#### 3.2.4. Mean diffusivity

Evidence largely favoured the null: systolic BP (β = −0.006, [−0.062, 0.048], BF₁₂ = 34.40) showed very strong evidence for the null, and diastolic BP (β = 0.036, [−0.015, 0.087], BF₁₂ = 15.17), BMI (β = 0.028, [−0.086, 0.144], BF₁₂ = 15.72), and HbA1c (β = −0.013, [−0.107, 0.074], BF₁₂ = 20.78) each showed strong evidence for the null. HDL cholesterol showed anecdotal evidence for a negative within-person association (β = −0.080, [−0.147, −0.013], BF₁₂ = 2.230).

### 3.3. Prospective prediction

Our prospective lagged model tested whether cardiometabolic indices at time *_t_* predict next-wave global brain structure at *_t + 1_* over and above the brain measure at *_t_*. Figure 4 shows the posterior distributions for associations between each incremental prediction from CMR to next wave global brain features.

**Figure 4.**
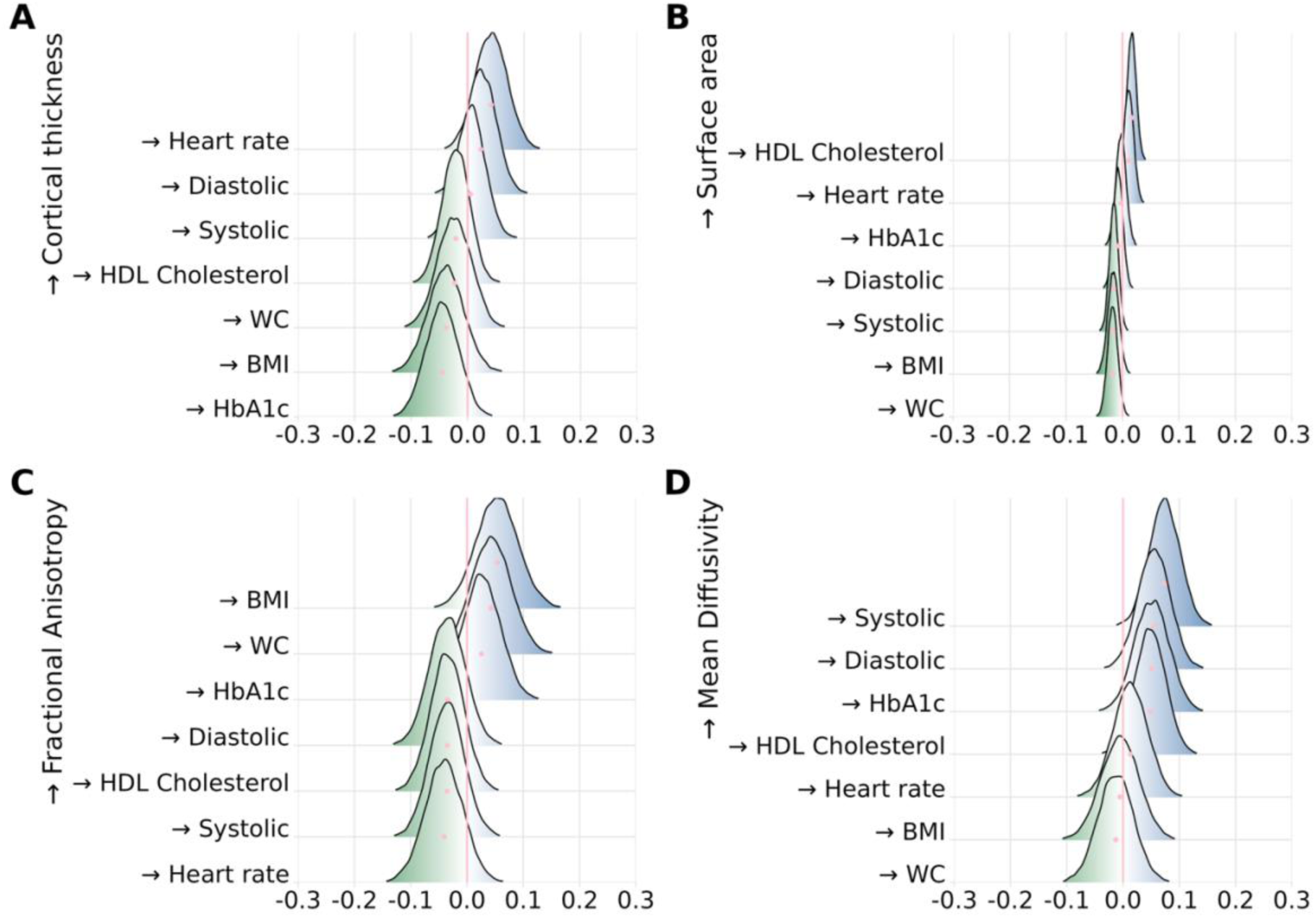
Lagged prospective associations between baseline CMRs and next-wave global brain structure. Posterior distributions of the standardised coefficients from the lagged model brain _t+1_ ∼ CMR _t_ + brain _t_ + Age + Sex + SES + (1|ID), estimating the incremental prediction of cardiometabolic status at time _t_ for (A) cortical thickness, (B) surface area, (C) fractional anisotropy, and (D) mean diffusivity at time *_t + 1_*, conditional on the prior brain measure. The → symbol denotes prospective prediction from CMR at time t to brain structure at the subsequent wave t+1, over and above prior brain level. Colour indicates direction (blue = positive; green = negative); distribution width reflects uncertainty. Abbreviations as in Figure 2.

#### 3.3.1. Cortical thickness

Across CMRs, evidence favoured the null: systolic BP (β = 0.005, [−0.046, 0.055], BF₁₂ = 38.16, very strong M1), diastolic BP (β = 0.023, [−0.031, 0.074], BF₁₂ = 25.05, strong M1), heart rate (β = 0.042, [−0.015, 0.097], BF₁₂ = 12.17, strong M1), WC (β = −0.023, [−0.081, 0.035], BF₁₂ = 24.11, strong M1), BMI (β = −0.038, [−0.101, 0.025], BF₁₂ = 15.27, strong M1), HbA1c (β = −0.045, [−0.101, 0.013], BF₁₂ = 11.60, strong M1), and HDL cholesterol (β = −0.021, [−0.070, 0.025], BF₁₂ = 28.98, strong M1); all providing strong to very strong evidence for no prospective association.

#### 3.3.2. Surface area

Findings consistently favoured the null. HbA1c showed extreme evidence for the null (β = −0.002, [−0.020, 0.016], BF₁₂ = 108.75). Diastolic BP (β = −0.008, [−0.024, 0.009], BF₁₂ = 77.32) and heart rate (β = 0.011, [−0.006, 0.028], BF₁₂ = 51.67) showed very strong evidence for the null. Systolic BP (β = −0.015, [−0.030, 0.001], BF₁₂ = 19.98), WC (β = −0.017, [−0.036, 0.000], BF₁₂ = 16.97), BMI (β = −0.016, [−0.036, 0.003], BF₁₂ = 26.15), and HDL cholesterol (β = 0.016, [0.001, 0.032], BF₁₂ = 15.00) all showed strong evidence for the null.

#### 3.3.3. Fractional anisotropy

All CMRs showed strong evidence for no prospective association, with the exception of BMI which showed moderate evidence for the null: systolic BP (β = −0.035, [−0.096, 0.025], BF₁₂ = 16.54), diastolic BP (β = −0.035, [−0.097, 0.028], BF₁₂ = 17.55), heart rate (β = −0.041, [−0.109, 0.026], BF₁₂ = 14.86), WC (β = 0.043, [−0.027, 0.111], BF₁₂ = 13.96), BMI (β = 0.054, [−0.020, 0.128], BF₁₂ = 9.657, moderate M1), HbA1c (β = 0.026, [−0.041, 0.091], BF₁₂ = 21.63), and HDL cholesterol (β = −0.035, [−0.094, 0.024], BF₁₂ = 16.73).

#### 3.3.4. Mean diffusivity

Most predictors favoured the null. Heart rate (β = 0.013, [−0.045, 0.075], BF₁₂ = 30.08), WC (β = −0.013, [−0.073, 0.051], BF₁₂ = 28.54), and BMI (β = −0.005, [−0.072, 0.060], BF₁₂ = 28.91) showed strong evidence for the null. Diastolic BP (β = 0.055, [−0.001, 0.112], BF₁₂ = 5.853), HbA1c (β = 0.051, [−0.007, 0.113], BF₁₂ = 7.959), and HDL cholesterol (β = 0.048, [−0.006, 0.103], BF₁₂ = 8.345) showed moderate evidence for the null. Systolic BP showed anecdotal evidence for the null (β = 0.074, [0.018, 0.126], BF₁₂ = 1.244).

### 3.4. Puberty adjusted models

Replacing age with pubertal status left the overall patterns of our findings unchanged. The full results are available in SI Table 8, with posterior distributions showing puberty-adjusted associations between CMRs and brain structure available in SI Figure 10. The sample distribution PDS scores split by sex are visualised in SI Figure 11.

## 4. Discussion

Across specifications, the dominant pattern was small, often null associations between cardiometabolic risk factors and global brain structure and microstructure in youth. In our primary multilevel model, most posteriors centred near zero for CT, SA, FA, and MD, with two exceptions: higher resting heart rate was related to higher MD, both across time points and strengthening across waves, and higher waist circumference showed a modest positive association with surface area. Outside these patterns, both main effects and time-interaction terms generally showed strong-to-extreme evidence for the null, indicating that any cross-sectional or time-varying associations, if present, are subtle.

When we focused on within-person change (Δbrain ∼ ΔCMR), the evidence again favoured the null across outcomes, indicating no co-movement of cardiometabolic and brain changes between adjacent visits. Likewise, in lagged prospective models that tested whether CMR at _t_ predicts brain at _t + 1_ over and above brain at _t_, Bayes Factors were strong-to-extreme for the null for nearly all predictor-outcome pairs. Importantly, these longitudinal tests span adjacent ABCD assessment waves separated by approximately two years, capturing relatively short-term developmental intervals; null effects therefore do not preclude the possibility of longer-lag associations emerging over extended follow-up; equally, the approximately two-year interval between waves may be too coarse to capture effects operating at shorter timescales, and finer-grained longitudinal designs with more frequent assessments may reveal associations that the current design is unable to detect. Taken together, these sensitivity analyses suggest that the few between-person associations observed in the primary models (e.g., WC-SA; heart-rate-MD) do not manifest as robust within-person co-variation.

To contextualise this interpretation, we note that within-person variability in cardiometabolic indices across waves was modest in our sample; mean within-person standard deviations ranged from approximately 0.15% for HbA1c to 8.0 mmHg for systolic blood pressure (see SI Table 2). Blood pressure and heart rate showed the largest absolute within-person variability yet the poorest temporal stability (systolic ICC = 0.40, diastolic ICC = 0.35, heart rate ICC = 0.43), consistent with the interpretation that single visit-based measurements of these indices are noisy proxies of habitual cardiovascular health. BMI showed the greatest stability (ICC = 0.77, within-person SD = 1.9 kg/m²), reflecting its well-established consistency as an anthropometric measure. This limited within-person variability may itself partly explain the null change-score findings, as small fluctuations in cardiometabolic indices over two-year intervals may be insufficient to drive detectable concurrent changes in global brain structure.

Two implications follow. First, the between-person signals, i.e., youths with higher resting heart rate have higher MD and youths with higher waist circumference have larger surface area, do not straightforwardly translate into within-person change or prospective effects once prior brain level is accounted for. This pattern fits a scenario where cardiometabolic status marks stable between-child differences (and/or shared correlates) more than it drives short-term global brain change. Relatedly, interpreting what constitutes a less favourable brain profile in adolescence is challenging, as cortical thickness follows predictable age-related trajectories and thinner cortex is not inherently adverse. The field also lacks consensus on how to distinguish normative maturation from delayed, accelerated, or adverse processes in this age range, underscoring the need for caution when interpreting structural differences.

Second, the convergence of strong-to-extreme evidence for the null across change-score and lagged models argues against sizeable direct effects of cardiometabolic indices on global brain metrics over the timepoints studied. If effects exist, they are likely small, region-specific, nonlinear, or contingent on maturational context, possibilities future work should probe. Finally, replacing age with pubertal development as a maturational covariate did not alter the overall conclusions, suggesting our findings are robust to maturational adjustment. In sum, across complementary modelling frames, the evidence indicates that global brain structure is largely insensitive to contemporaneous cardiometabolic variation in this developmental window, with heart-rate-MD (higher MD) and WC-SA (larger surface area) as the most consistent, yet still modest, exceptions.

Population work likewise finds that body-fat indices relate to brain metrics whereas other cardiometabolic markers (blood pressure, glucose/insulin, lipids) often show little association, mirroring our findings^21^. Although BMI and waist circumference are correlated measures of adiposity, they showed divergent patterns in the current study – with waist circumference showing a modest positive association with surface area while BMI showed no clear association with any brain metric. This may reflect the fact that BMI incorporates height, which changes substantially across the age range studied, whereas waist circumference does not, potentially capturing different aspects of body composition with distinct neural correlates; but this is speculative.

The association we observe between resting heart rate and elevated mean diffusivity is compatible with literature positioning heart rate as a coarse proxy for cardiorespiratory fitness, whereby higher fitness is associated with healthier WMM in children and adolescents^50,51^, although, this remains speculative in the current context, and future work directly incorporating objective measures of cardiorespiratory fitness available in the ABCD dataset would be valuable for testing this more rigorously. More broadly, these associations (and our largely null change-score and lagged effects) illustrate a key longitudinal principle: between-person differences seldom guarantee within-person sensitivity at the same timescale, because stable trait variance can drive cross-sectional signals that do not translate into time-ordered within-child effects^52,53^.

We found no blood pressure associations with global brain metrics across waves, consistent with prior reports of attenuated diastolic effects after early-life adjustment^24^ and extending them by showing no detectable relations with cortical or diffusion measures. This aligns with findings from the ABCD Study showing that when individual cardiometabolic markers are modelled separately, associations with brain structure and neurocognitive outcomes are largely null^25^.

Several mechanisms may explain why robust between-child associations rarely manifest as short-term within-child co-variation, most notably the influence of shared liabilities that shape both cardiometabolic status and brain development over longer timescales. For example, multivariate brain features and impulsivity at baseline predict subsequent rapid weight-gain trajectories^54^; prenatal metabolic exposures (e.g., gestational diabetes) are associated with lower global and regional grey matter volume and partially mediate later adiposity^55^; youth with type 2 diabetes show more pronounced subcortical and white matter differences than BMI-matched peers^56^; and socioeconomic, behavioural, and genetic factors, are independently associated with both cardiometabolic health and brain variation^57,58^.

Together, these findings situate our modest heart-rate-MD and WC-SA associations within a framework of shared developmental and contextual influences, while reinforcing that short-term, global-scale within-child effects of common cardiometabolic markers are likely small during early-to-mid adolescence. These interpretations remain tentative. More studies are needed to fill critical gaps, and future research should make use of larger samples with better quality integrated imaging-clinical-biological mediators, and possibly intervention studies (e.g., weight loss programs) to assess direction of effects. Future research could also build on the present findings by incorporating clinically informed cardiometabolic risk stratification (e.g., risk vs. healthy groupings) to examine whether threshold effects or categorical risk status show differential associations with brain development trajectories.

### 4.1. Strengths and limitations

In terms of strengths, we leverage the latest ABCD 6.0 release to assemble a large, multi-site, three-wave dataset with harmonised imaging and rigorous quality control, random-effects modelling that respects within-participant dependency, principled handling of age-time collinearity (age centred within timepoint), and complementary specifications (time-interaction, within-person change, and lagged models). Bayesian estimation enables calibrated interpretation of null results, which is essential given widespread small effects in youth neuroimaging. Sensitivity analyses replacing chronological age with pubertal development status led to similar conclusions, increasing confidence that results are not artefacts of maturational adjustment strategy.

Limitations include missingness and irregular longitudinal coverage of the sample. Of the 3527 participants, many youth contributed only a single wave, and only 782 participants contributing repeated measurements (658 across two waves and 124 across all three waves), reducing power for change and lagged models and increasing vulnerability to selective attrition. Formal attrition analyses indicated that participants contributing longitudinal data did not differ substantially from cross-sectional-only participants on most key variables, though age, systolic blood pressure, and HbA1c showed small but significant differences.

Among participants with repeated measurements, those completing all three waves showed mild health-related retention bias, being slightly younger with lower BMI and higher HDL cholesterol, suggesting within-person estimates may be modestly conservative. Results of attrition analyses are provided in SI Figure 1 and SI Table 2. Next, despite longCombat harmonisation and extensive QA, residual confounding remains a limitation. Variables including sleep, diet, and genetic liability remain important unmeasured sources of confounding that future work should address. However, the use of Bayesian regularising priors, random effects for subject ID, and convergent evidence across three complementary model specifications provide some protection against spurious findings driven by unmeasured confounders.

Although we probed interactions with time and used change and lagged models, non-linear developmental effects, thresholds, and sex-specific patterns may still require more flexible models and additional waves. Furthermore, the focus on global brain measures does not capture spatially specific associations that may exist at the regional level; future work should examine whether cardiometabolic risk indices relate differentially to regional cortical and white matter development during adolescence. Our lagged models did not investigate potential bidirectional effects, i.e., whether brain measures at one wave predict CMR at a later wave. While we acknowledge this as a meaningful future direction and note that the reverse direction is theoretically plausible given the reciprocal brain-body connection, the current study was specifically designed to test whether cardiometabolic health predicts brain development, and the reverse direction constitutes a distinct research question that we believe warrants dedicated investigation in future work. Additionally, raw change scores carry measurement error from both time points, and while the Bayesian multilevel framework with regularising priors provides a conservative estimation approach, future work with more repeated observations per participant could more formally account for measurement error through reliable change indices or individual growth curve modelling.

Visit-based measurements of resting heart rate and blood pressure may not optimally reflect habitual cardiometabolic health, as transient elevations due to visit-related stress or white coat effects could attenuate true associations with brain structure. Future work leveraging more ecologically valid approaches, such as wearable-based continuous monitoring data available in a subset of ABCD participants, may reveal associations that single visit-based measurements are insufficiently sensitive to detect. Finally, generalisability is constrained by both sample composition and data availability. The analytic sample represents a subset of the full ABCD cohort, primarily due to the requirement for overlapping CMR and MRI data at the same assessment wave and the limited availability of blood assay measures across the broader cohort. Beyond the ABCD cohort itself, results may not extend to clinical populations or contexts with markedly different sociocultural and environmental exposures.

## 5. Conclusion

Across three waves spanning late childhood to late adolescence, we found higher resting heart rate associated with higher mean diffusivity (strengthening over time), and higher waist circumference associated with larger surface area. Effect sizes were modest, and between-person associations did not consistently translate into within-person co-variation or prospective prediction beyond prior brain level, though these longitudinal inferences were informed by a relatively small subsample of participants with repeated measurements (n = 782), and should be interpreted with appropriate caution. Within-person variability in cardiometabolic indices was modest across the study period – mean within-person standard deviations ranged from 0.15% for HbA1c to 8.0 mmHg for systolic blood pressure – which may itself partly limit the sensitivity of within-person analyses to detect concurrent brain change over two-year intervals. Taken together, these findings suggest that, in early-mid adolescence, global brain architecture shows limited sensitivity to the degree of cardiometabolic variation observed in this sample over this timeframe. Efforts to improve cardiometabolic health in youth remain vital for broader medical and cognitive outcomes, but neuroscience investigations may need larger longitudinal samples, more precise and ecologically valid phenotyping, longer follow-up, and causal designs to determine how cardiometabolic risk impacts the developing brain.

## 6. Acknowledgement

Data used in the preparation of this article were obtained from the <u>Adolescent Brain</u> <u>Cognitive Development™ (ABCD) Study</u>, held in the <u>NIH Brain Development Cohorts Data</u> <u>Sharing Platform</u>. This is a multisite, longitudinal study designed to recruit more than 10,000 children aged 9-10 and follow them over 10 years into early adulthood. The ABCD Study® is supported by the National Institutes of Health and additional federal partners under award numbers: U01DA041048, U01DA050989, U01DA051016, U01DA041022, U01DA051018, U01DA051037, U01DA050987, U01DA041174, U01DA041106, U01DA041117, U01DA041028, U01DA041134, U01DA050988, U01DA051039, U01DA041156, U01DA041025, U01DA041120, U01DA051038, U01DA041148, U01DA041093, U01DA041089, U24DA041123, U24DA041147. A full list of supporters is available at <u>Federal Partners - ABCD Study</u>. ABCD Consortium investigators designed and implemented the study and/or provided data but did not necessarily participate in the analysis or writing of this report. This manuscript reflects the views of the authors and may not reflect the opinions or views of the NIH or ABCD Consortium investigators.

Data was handled inside Service for Sensitive Data (TSD), a platform owned by the University of Oslo, operated, and developed by the TSD service group at the University of Oslo IT-Department (USIT). Computations were also performed using resources provided by UNINETT Sigma2 – the National Infrastructure for High Performance Computing and Data Storage in Norway (NS9666S).

## 7. Funding

This research is supported by the Research Council of Norway (#288083, #323951, #300767), and the South-Eastern Norway Regional Health Authority (#2021070, #2023012, #500189).

## 8. Conflict of interest statement

The authors declare that they have no conflict of interest.

## 9. Data availability statement

The data that support the findings of this study are available from the Adolescent Brain Cognitive Development™ (ABCD) Study via the NIH Brain Development Cohorts (NBDC) Data Hub. The specific dataset used was ABCD Study Release 6.0 (DOI: https://doi.org/10.82525/jy7n-g441). Access to these data is subject to approval through the NIH Data Use Certification (DUC) process, and the data are available to qualified researchers upon reasonable request and with appropriate authorization via the NBDC Data Hub (https://nbdc-datahub.org).

## 10. CRediT author contribution statement

**Beck, Dani**: Writing – review & editing, Visualisation, Methodology, Formal analysis, Data curation, Conceptualisation. **Westlye, Lars T.**: Writing – review & editing, Funding acquisition, Conceptualisation. **Tamnes, Christian K.**: Writing – review & editing, Project administration, Supervision, Funding acquisition, Conceptualisation.

## Notes

### Competing Interest Statement

The authors have declared no competing interest.

### Author Declarations

The current study uses data from the Adolescent Brain Cognitive Development (ABCD) Study. The Institutional Review Board (IRB) at the University of California, San Diego, approved all aspects of the ABCD Study. The current study was conducted in line with the Declaration of Helsinki and was approved by the Norwegian Regional Committee for Medical and Health Research Ethics (REK 2019/943).

### Summary of Updates

Socioeconomic status was included as a covariate for all analyses.

