## Supplementary Material for "Cardiometabolic risk and structural brain development in a large community-based U.S. cohort"

**SI Section 1.** Quality assurance.

The initial N = ~11,800 included in the study were drastically reduced largely due to limited cardiometabolic risk (CMR) data, which we chose to treat as a sub-sample rather than imputing data or using available data from sessions that did not match MRI data collection sessions or vice versa.

Global cortical thickness and surface area were selected as complementary cortical measures reflecting distinct neurodevelopmental processes: cortical thickness indexes age-related maturational thinning during adolescence, while surface area is largely determined earlier in development and shows relative stability across this period. Fractional anisotropy and mean diffusivity were selected as the most widely used and interpretable diffusion tensor imaging indices of white matter microstructural organisation in developmental neuroimaging, capturing complementary aspects of tissue organisation. Global rather than regional brain measures were selected due to their ability to provide a broad characterisation of structural and microstructural properties across the brain simultaneously, allowing us to assess whether cardiometabolic health is associated with brain integrity at a macro level.

For brain measures, quality control procedures followed a standard protocol described in Hagler et al. (2019). Briefly, participants with excessive head motion or poor data quality were excluded from the curated data release by the ABCD Study team. The imported tabulated MRI data initially included 30,276 obs for cortical thickness (CT; data structure smri_thick_cdk_mean) and cortical surface area (SA; data structure smri_area_cdk_total), and 29,165 obs for fractional anisotropy (FA; data structure mri_y_dti_fa_is_at) and mean diffusivity (MD; data structure mri_y_dti_md_is_at). Next, additional quality assurance was carried out following extraction of data using the recommendations for data cleaning provided by the ABCD Study team (using data structure abcd_imgincl01). Following this step, the obs for CT and SA were 29,594 and the obs for FA and MD were 25,949, which were merged together to form a combined MRI data frame of 25,949 obs, which was further reduced to obs = 15,712 (ses-02A N = 6,838, ses-04A N = 5,459, ses-06A N = 3,415) when removing ses-00A (baseline) to match CMR data that had no blood assay measures from this session (discussed below). Following these recommended steps, the data were then separately used for long.combat, described in SI Section 2 above, before being merged with the CMR data.

The cardiometabolic measures examined – BMI, waist circumference, systolic and diastolic blood pressure, resting heart rate, HbA1c, and HDL cholesterol – were selected because they represent the three primary cardiometabolic domains (anthropometric, cardiovascular, and metabolic), are routinely collected in paediatric clinical settings, and have established links to health outcomes in youth. Selection was also guided by data availability: additional blood assay measures were initially considered but excluded due to high missingness rates across waves, reflecting a deliberate decision to prioritise data quality and completeness over breadth of measures.

For cardiometabolic risk factors (CMRs); starting with measures from the blood draw data (including Haemoglobin A1c (HbA1c), High-Density Lipoprotein (HDL) Cholesterol): from the initial ~11,800 participants enrolled in the ABCD Study, only 9,024 obs in data frame *ph_y_bld.tsv* had Cholesterol (not used in the current study), HDL cholesterol, and HbA1c data from all the available sessions (ses-02A N = 1407, ses-03A N = 828, ses-04A N = 2501, ses-05A N = 1805, ses-06A N = 2483). This is reduced to 6,391 obs when filtering the sessions to match MRI sessions (ses-02A N = 1407, ses-04A N = 2501, and ses-06A N = 2483). Next, participants that had missing data for all three variables used in this data frame (HDL Cholesterol, Cholesterol, HbA1c) were removed, resulting in 5201 obs including ses-02A N = 1133, ses-04A N = 1748, and ses-06A N = 2320. Total missingness from the cholesterol variable was still ~20% and thus we chose to omit this variable from downstream analysis.

Next, the anthropometric data was imported (*ph_y_anthr.tsv*) and included 58,581 obs (ses-00A N = 11,861, ses-01A N = 11,168, ses-02A N = 9,201, ses-03A N = 4,095, ses-04A N = 8,964, ses-05A N = 8,297, ses-06A N = 4,995). This was reduced to 23,160 obs when filtering the sessions to match MRI sessions (ses-02A N = 9,201, ses-04A N = 8,964, and ses-06A N = 4,995). From this data frame, body-mass index (BMI) and waist circumference (WC) were used for downstream analyses.	Next, the blood pressure data was imported (*ph_y_bp.tsv*) and initially included 20,733 obs (ses-02A N = 4,788, ses-03A N = 1,639, ses-04A N = 7,347, ses-05A N = 2,239, ses-06A N = 4,720). This was reduced to 16,855 obs when filtering the sessions to match MRI sessions (ses-02A N = 4,788, ses-04A N = 7,347, and ses-06A N = 4,720). From this data frame, systolic blood pressure (SBP), diastolic blood pressure (DBP), as well as resting heart rate were used for downstream analyses.

Quality assurance procedures of cardiometabolic risk data included, for BMI, removal of all measures below and equal to a BMI of 10 and above and equal to a BMI of 50 due to the likelihood of these representing measurement errors in the data set. WC was cleaned by removing all measures below and equal to a WC of 15 and above and equal to a WC of 150. As a further measure of quality assurance, we checked the sample height at TP2 versus TP3 to ensure no participant was taller in TP2 than TP3. This was also done for TP3-TP4. During this step, no participant was flagged for removal. This cleaned data frame was merged with the blood pressure measures of systolic, diastolic, and heart rate, and blood assay measures of HbA1c, total and HDL cholesterol, which were all checked for outliers with manual inspection of histograms and outlier identification and removal of values beyond ± 5 SD. Three participants with implausible systolic and diastolic BP data were removed from the data frame, with no measures of heart rate removed. Four participants were removed with implausible HbA1c levels, with one removed from HDL cholesterol. The three data frames including blood assay measures, blood pressure measures, and anthropometric measures were merged together to form the final data frame pertaining to cardiometabolic risk factors (obs = 5,010: ses-02A N = 1,073, ses-04A N = 1,680, and ses-06A N = 2,257). SI Figures 6 and 7 show boxplots before and after quality assurance procedures for each CMR measure.

Due to the large discrepancy between the CMR and MRI data frames – and thus potentially large amount of missing data in the final data structure – the MRI data frame was merged into (and reduced to) the CMR data frame, to form a sub-sample for the current study. The sample size remained the same as reported in the paragraph above (obs = 5,010: ses-02A N = 1,073, ses-04A N = 1,680, and ses-06A N = 2,257).

Finally, to minimise confounding effects from complex family-related factors, unrelated participants were chosen by randomly selecting an individual from each family ID, subsequently excluding any siblings. The final sample consisted of 4,433 obs from 3,527 unique participants (ses-02A N = 970, ses-04A N = 1,493, ses-06A N = 1,970).

| **SI Table 1**. Demographic and neuroimaging characteristics of the full ABCD Study sample following quality assurance procedures and the analytic subsample used in the current study. | | |
| --- | --- | --- |
|  | **Full ABCD sample** | **Analytic subsample** |
|  | N = 11,192 | N = 3,527 |
| **Age (years)** | 12.12 (0.76) | 12.06 (0.72) |
| **Sex** |  |  |
| **Female** | 5,315 (47%) | 1,666 (47%) |
| **Male** | 5,877 (53%) | 1,861 (53%) |
| **Ethnicity** |  |  |
| **Asian** | 214 (1.9%) | 61 (1.7%) |
| **Black** | 1,636 (15%) | 458 (13%) |
| **Hispanic** | 2,264 (20%) | 708 (20%) |
| **Other** | 1,131 (10%) | 334 (9.5%) |
| **White** | 5,945 (53%) | 1,966 (56%) |
| **SES** | 3.75 (1.26) | 3.78 (1.26) |
| **PDS** | 2.14 (0.70) | 2.14 (0.70) |
| **BMI (kg/m^2^)** | 20.70 (4.82) | 20.70 (4.82) |
| **WC (cm)** | 72.86 (12.21) | 72.86 (12.21) |
| **Systolic (mmHg)** | 103.48 (10.72) | 103.48 (10.72) |
| **Diastolic (mmHg)** | 60.41 (8.50) | 60.41 (8.50) |
| **Heart rate (bpm)** | 78.06 (11.07) | 78.06 (11.07) |
| **HDL Cholesterol (mg/dL)** | 56.62 (13.17) | 56.62 (13.17) |
| **Hba1c (%)** | 5.14 (0.29) | 5.14 (0.29) |
| **FA (ratio)** | 0.51 (0.02) | 0.51 (0.02) |
| **MD (mm^2^/s)** | 0.79 (0.02) | 0.79 (0.02) |
| **Thickness (mm)** | 2.69 (0.08) | 2.69 (0.08) |
| **Area (mm^2^)** | 190,458.44 (18,456.25) | 190,458.44 (18,456.25) |

**Note**: Full ABCD sample: all participants with available data following ABCD Study quality assurance procedures (N = 11,192); Analytic subsample: participants with overlapping cardiometabolic and MRI data at the same assessment wave retained for primary analyses (N = 3,527). Values represent mean (SD) for continuous variables and n (%) for categorical variables. Comparison is limited to variables available across both samples – blood pressure, HbA1c, and HDL cholesterol are not available for the full ABCD cohort outside the analytic subsample and are therefore not included. The primary driver of exclusion from the analytic sample was the requirement for overlapping CMR and MRI data at the same assessment wave, and in particular the limited availability of blood assay measures across the full cohort. Waves refer to the two-, four-, and six-year follow-up assessment sessions (ses-02A, ses-04A, ses-06A). BMI: body-mass index (kg/m²); WC: waist circumference (cm); FA: fractional anisotropy (dimensionless); MD: mean diffusivity (mm²/s); CT: cortical thickness (mm); SA: surface area (mm²); SES: composite socioeconomic status score incorporating household income and parental education; PDS: Pubertal Development Scale caregiver-reported sum score (higher scores reflect more advanced pubertal development).

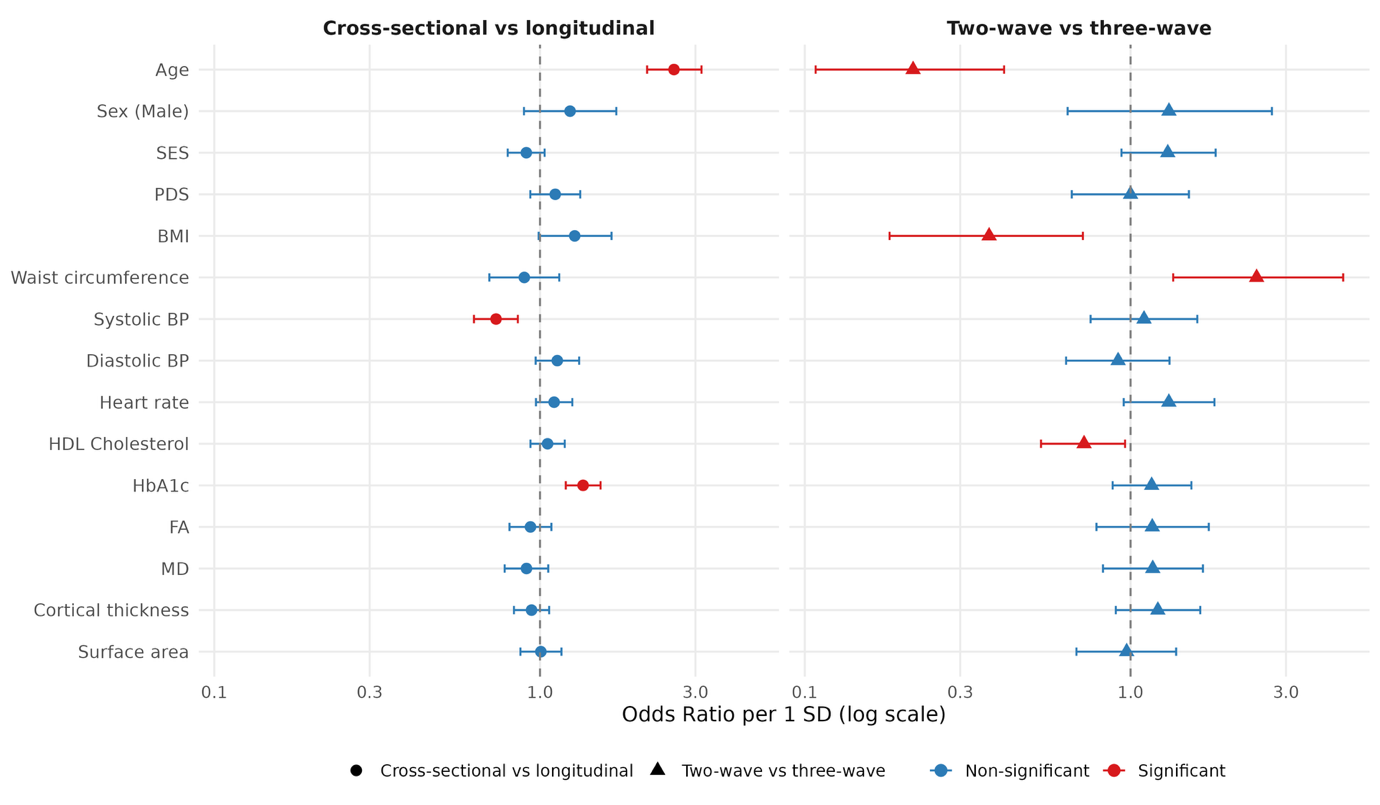

**SI Figure 1**. Odds ratios (ORs) from two logistic regression models predicting participation pattern, with all predictors standardised (per 1 SD) to allow comparison across variables. Left panel: cross-sectional only vs. any longitudinal participation (n = 3,527); right panel: two-wave vs. three-wave retention among longitudinal participants (n = 782). Points indicate OR estimates; horizontal bars indicate 95% confidence intervals. Red = statistically significant (p < .05); blue = non-significant. In the primary attrition model, age, systolic blood pressure, and HbA1c were the only significant predictors of longitudinal participation, with SES, BMI, waist circumference, brain MRI measures, and most other cardiometabolic variables all non-significant. In the secondary retention model, age, BMI, waist circumference, and HDL cholesterol were significant, suggesting mild health-related retention bias in the three-wave subsample whereby slightly healthier participants were more likely to complete all three waves.

**SI Table 2**. Intraclass correlation coefficients (ICC2, two-way random effects model, single measures) and mean within-person standard deviations (SD) for each cardiometabolic risk measure, calculated for participants with repeated measurements (n = 782). Higher ICC values indicate greater temporal stability across waves.

| **Variable** | **ICC** | **Lower 95** | **Upper 95** | **Count** | **Within-person (mean)** | **Within-person (SD)** |
| --- | --- | --- | --- | --- | --- | --- |
| BMI | 0.767 | 0.503 | 0.876 | 122 | 1.921 | 1.585 |
| WC | 0.653 | 0.466 | 0.771 | 123 | 6.139 | 4.45 |
| Systolic | 0.39 | 0.191 | 0.553 | 124 | 8.006 | 5.655 |
| Diastolic | 0.348 | 0.215 | 0.476 | 124 | 5.837 | 4.591 |
| Heart rate | 0.439 | 0.308 | 0.558 | 124 | 7.283 | 5.573 |
| HDL Cholesterol | 0.557 | 0.428 | 0.671 | 90 | 6.62 | 5.292 |
| Hba1c | 0.614 | 0.346 | 0.769 | 91 | 0.153 | 0.111 |

**SI Section 2**. MRI acquisition and processing.

For T1-weighted MRI acquisitions on Siemens and Philips 3T scanners, a matrix of 256 × 256, 176 slices (Siemens) and 225 slices (Philips) with a field of view (FOV) of 256 × 256, echo time (TE)/repetition time (TR) (ms) of 2.88/2500 (Siemens) and 2.9/6.31 (Philips), flip angle of 8°. On GE scanners, the same matrix and FOV were used with TE/TR (ms) of 2/2500 and a flip angle of 8°. The spatial resolution was consistent at 1.0 × 1.0 × 1.0 mm across all three platforms. The dMRI acquisition (1.7 mm isotropic) uses multiband EPI with slice acceleration factor 3 and includes 96 diffusion directions, seven b = 0 frames, and four b-values (6 directions with b = 500 s/mm^2^, 15 directions with b = 1000 s/mm^2^, 15 directions with b = 2000 s/mm^2^, and 60 directions with b = 3000 s/mm^2^).

For T1-weighted MRI data, cortical surface reconstruction and subcortical segmentation was performed with FreeSurfer v7.1.1 ^2,3^. This processing includes motion correction and averaging ^4^, removal of non-brain tissue (Ségonne et al., 2004), automated Talairach transformation, segmentation of the subcortical white matter and deep gray matter volumetric structures ^3,5^, intensity normalisation ^6^, tessellation of the gray matter white matter boundary, automated topology correction ^7,8^, and surface deformation ^2^. From the ABCD Data Repository, we obtained tabulated data of total cortical surface area and thickness from the following data structures: mri_y_smr_thk_dsk, mri_y_smr_area_dsk.

For DTI data, processing was carried out using AtlasTrack, a probabilistic atlas-based method for automated segmentation of white matter fibre tracts (Hagler et al., 2009). A detailed pipeline can be found in Hagler et al. (2019). Briefly, diffusion tensor parameters are calculated using a standard, linear estimation approach with log-transformed diffusion-weighted (DW) signals ^11^, whereby two tensor models are fitted. In the first DTI model fit (DTI inner shell, or DTI_IS_), frames with b > 1000 s/mm^2^ are excluded from tensor fitting (leaving 6 directions at b = 500 s/mm^2^ and 15 directions at b = 1000 s/mm^2^) so that the derived diffusivity measures better correspond to those from traditional, single-b-value acquisitions. In the second DTI model fit (DTI full shell, or DTI_FS_), all gradient strengths/shells (6 directions at b = 500 s/mm^2^, 15 directions at b = 1000 s/mm^2^, 15 directions at b = 2000 s/mm^2^, and 60 directions at b = 3000 s/mm^2^) are included. For the current study, only inner shell tissue properties of functional anisotropy (FA) and mean (MD) were extracted for total (global) scores of each DTI feature. This involved obtaining the following tabulated data structures from the ABCD Data Repository: mri_y_dti_fa_is_at, mri_y_dti_md_is_at.csv. Quality assurance procedures carried out on all outlined brain features for each modality are outlined in SI Section 3 below.

**SI Section 3**. Long Combat.

The ABCD Study data is collected from 21 different sites in the U.S, and children are scanned using thirty-four different magnetic resonance imaging (MRI) scanners from three manufacturing brands (3-T Siemens Prisma, General Electric 750 or Phillips). Due to the technical variability of using multiple scanners, noise and bias can be introduced into the estimation of biological features of interest. Longitudinal ComBat is a powerful harmonisation procedure for longitudinal datasets and controls for type I error better than unharmonised data with scanner included as a covariate ^12^. In the current study, LongCombat was implemented on two-year, four-year, and six-year follow-up observations (obs) of cortical thickness, surface area, fractional anisotropy, and mean diffusivity data from the ABCD Study cohort (release 6.0), with each MRI modality being harmonised separately, including covariates of age, sex, timepoint, and socioeconomic status in the design matrix, following recommendations from Beer et al. (2020) and addressing potential demographic differences between ABCD Study sites. Scanner ID was used as the site variable, thereby also capturing head-coil differences between sites. SI Figure 1 shows MRI data from each modality for selected global features prior to and post harmonisation.

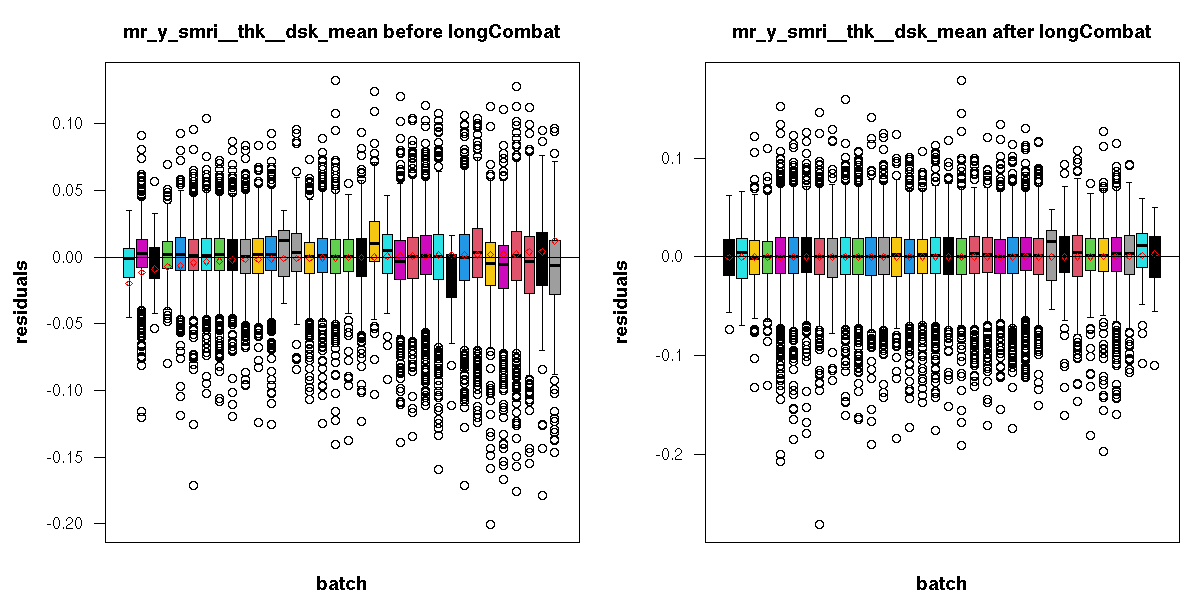

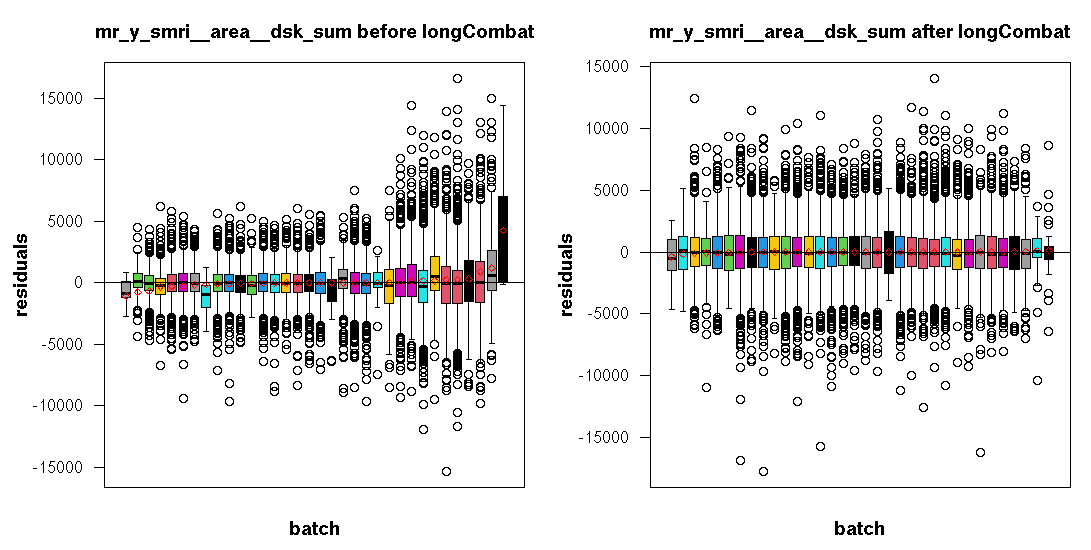

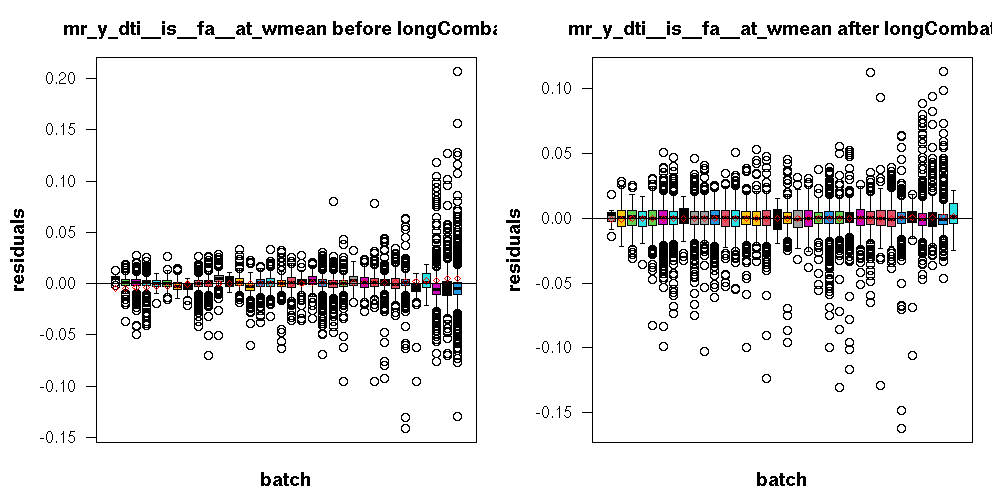

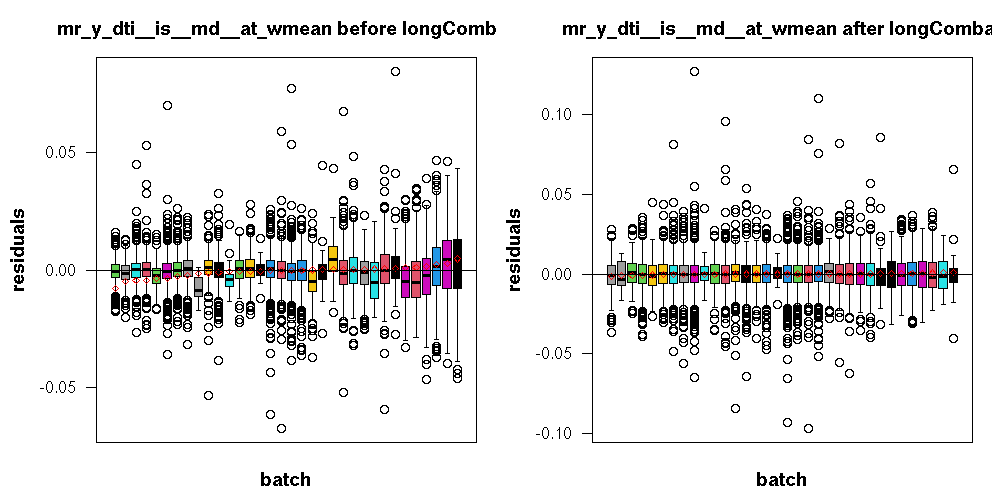

**SI Figure 2**. Showing harmonisation of scanner effects using long.combat for selected global brain MRI features used in the study.

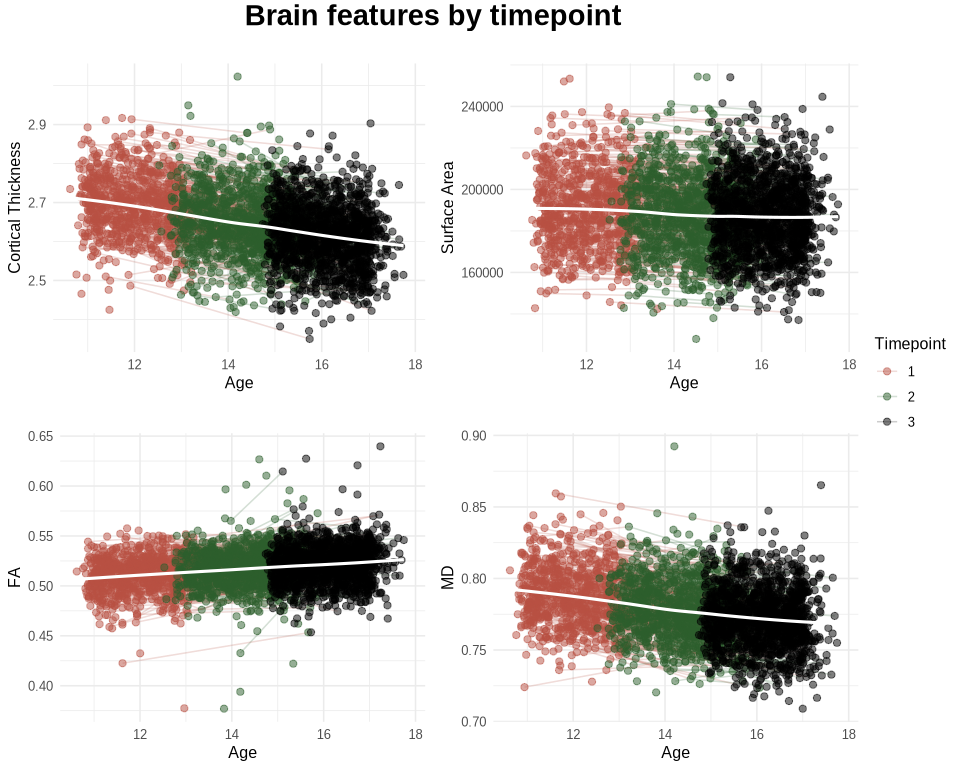

**SI Figure 3**. Age patterns for the study sample. Showing timepoint 1, 2, and 3 (year two, four, and six) of the ABCD Study for cortical thickness, surface area, fractional anisotropy (FA), mean diffusivity (MD).

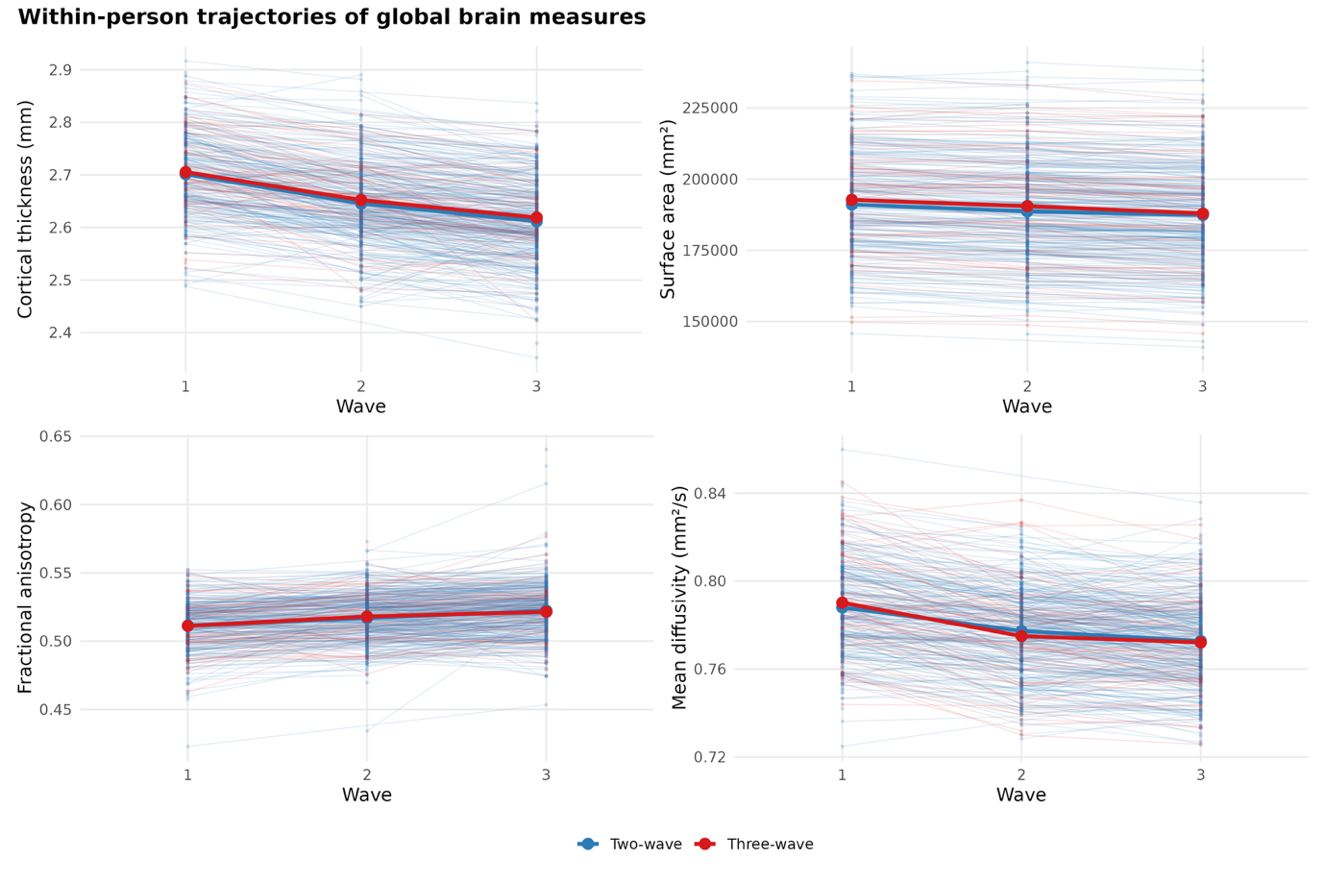

**SI Figure 4**. Within-person trajectories of global brain measures across study waves for participants contributing two waves (blue) and three waves (red) of data. Each line represents one participant; bold lines indicate group means. Figures are shown separately for cortical thickness, surface area, fractional anisotropy, and mean diffusivity.

### **SI Table 3**. Definitions of normal and elevated blood pressure, and hypertension among children and adolescents according to the 2017 American Academy of Paediatrics (AAP) Clinical Practice Guideline^13^.

| **Blood pressure category^a^** | **Children and adolescents aged 1–12 years** | **Adolescents aged ≥13 years** |
| --- | --- | --- |
| Normal blood pressure | Systolic BP and Diastolic BP <90th percentile^b^ | Systolic BP <120 mm Hg and Diastolic BP <80 mm Hg |
| Elevated blood pressure^c^ | Systolic BP and/or Diastolic BP ≥90th to <95th percentile or Systolic BP ≥120 mm Hg and Diastolic BP <80 mm Hg to the 95th percentile | Systolic BP 120–129 mm Hg and Diastolic BP <80 mm Hg |
| Stage 1 hypertension | Systolic BP and/or Diastolic BP ≥95th percentile to <95th percentile + 12 mm Hg or SBP 130–139 mm Hg and/or Diastolic BP 80–89 mm Hg | Systolic BP 130–139 mm Hg and/or Diastolic BP 80–89 mm Hg |
| Stage 2 hypertension | ≥95th percentile + 12 mm Hg or Systolic BP ≥140 mm Hg and/or Diastolic BP ≥90 mm Hg | Systolic BP ≥140 mm Hg and/or Diastolic BP ≥90 mm Hg |

Abbreviations: BP, blood pressure. ^a^Definitions were obtained from the 2017 AAP Clinical Practice Guideline.

^b^Percentiles were determined using age-, sex-, and height-specific percentile tables from the 2017 AAP Clinical Practice Guideline. ^c^High blood pressure was defined as having either elevated blood pressure or hypertension.

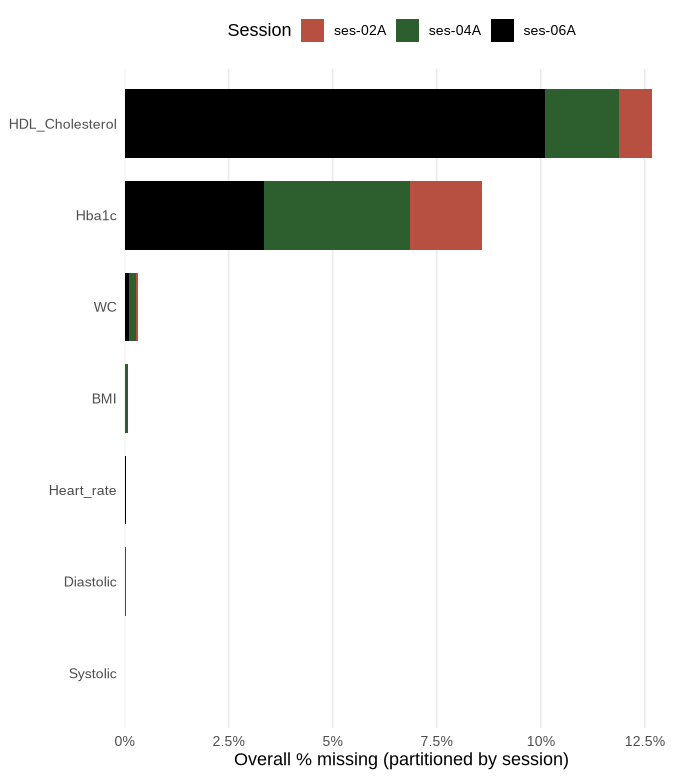

**SI Figure 5.** Percentage of missing data for each cardiometabolic risk variable within the final quality-assured CMR analytic sample (total observations = 5,010; ses-02A N = 1,073, ses-04A N = 1,680, ses-06A N = 2,257). This sample was formed by merging three separate data frames – blood assay measures (HbA1c, HDL cholesterol), blood pressure measures (systolic BP, diastolic BP, heart rate), and anthropometric measures (BMI, waist circumference) – following rigorous quality assurance procedures applied to each data frame independently (see SI Section 1). Residual missingness reflects the fact that not all participants had data available across all three data frames at each session. Bar length represents total missingness percentage; colours represent the proportion contributed by each session (ses-02A, ses-04A, ses-06A).

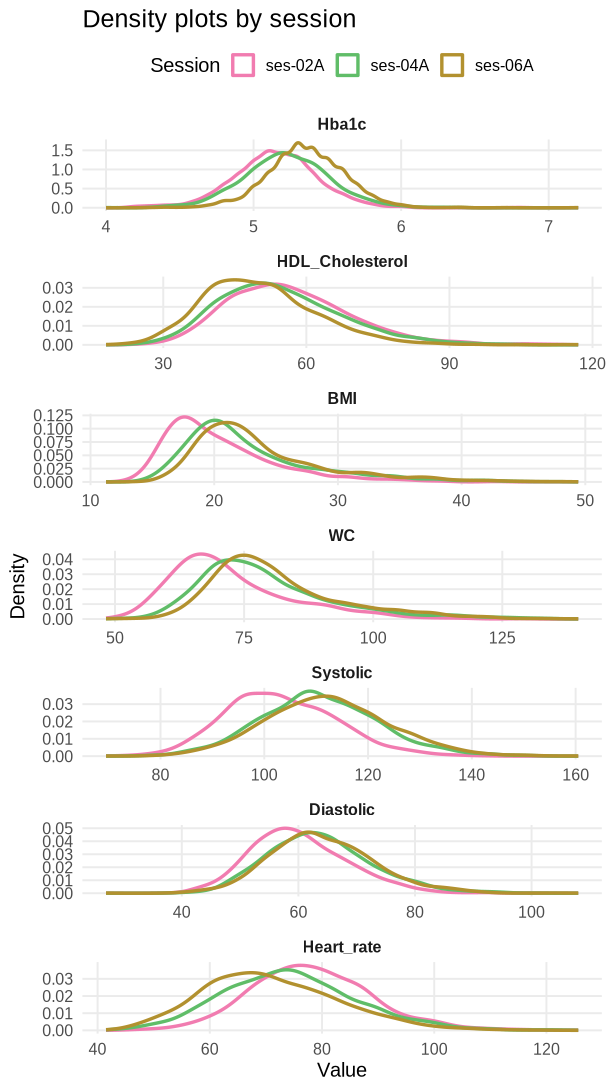

**SI Figure 6**. Density plots showing the distribution of each cardiometabolic risk measure across the three study waves (ses-02A, ses-04A, ses-06A). Note that clinically relevant thresholds vary by age and sex for several measures (BMI, blood pressure); relevant clinical definitions for all measures are provided in the Methods section 2.3 and SI Table 3.

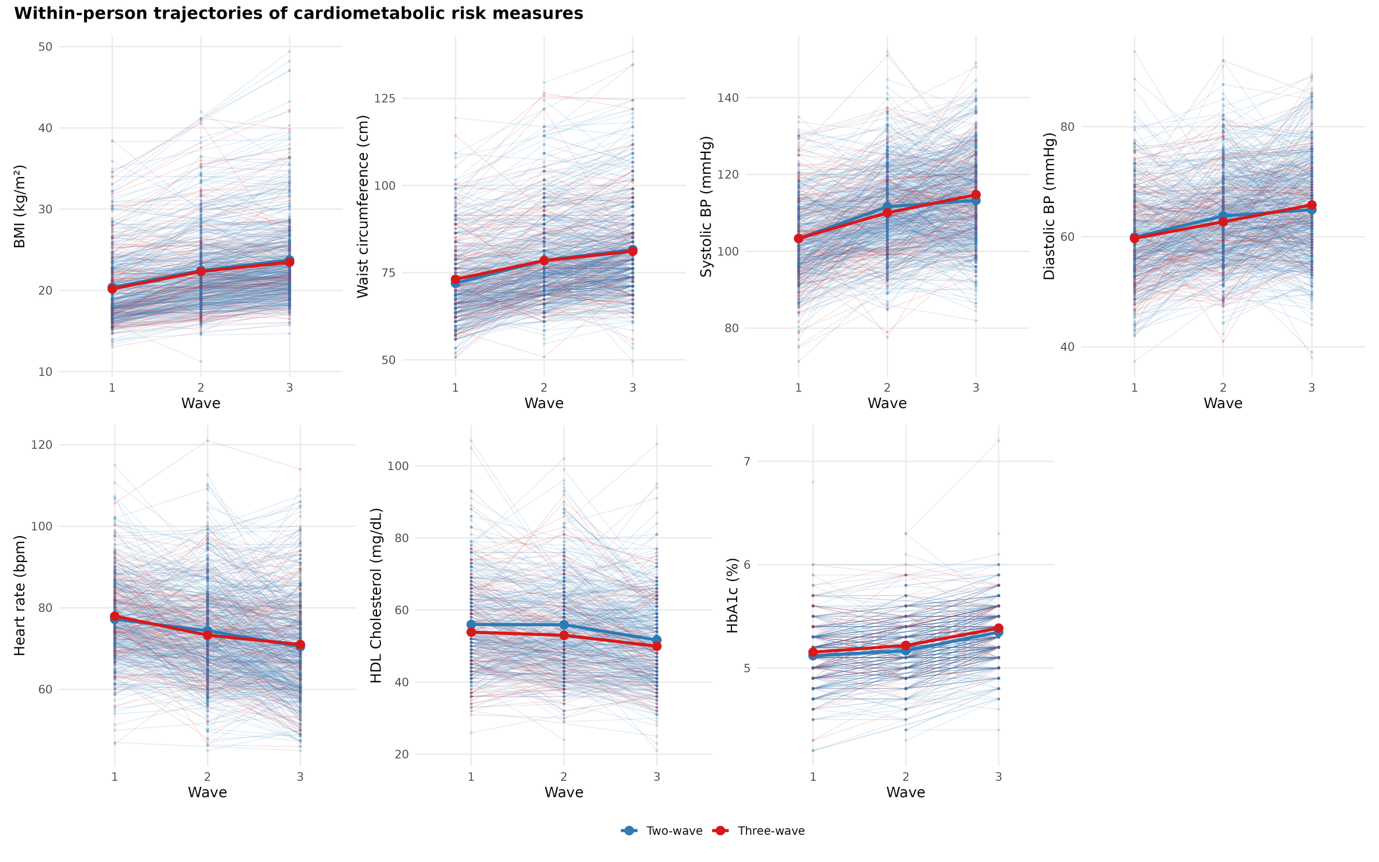

**SI Figure 7**. Within-person trajectories of cardiometabolic risk measures across study waves for participants contributing two waves (blue) and three waves (red) of data. Each line represents one participant; bold lines indicate group means. Figures are shown separately for BMI, waist circumference, systolic blood pressure, diastolic blood pressure, resting heart rate, HDL cholesterol, and HbA1c.

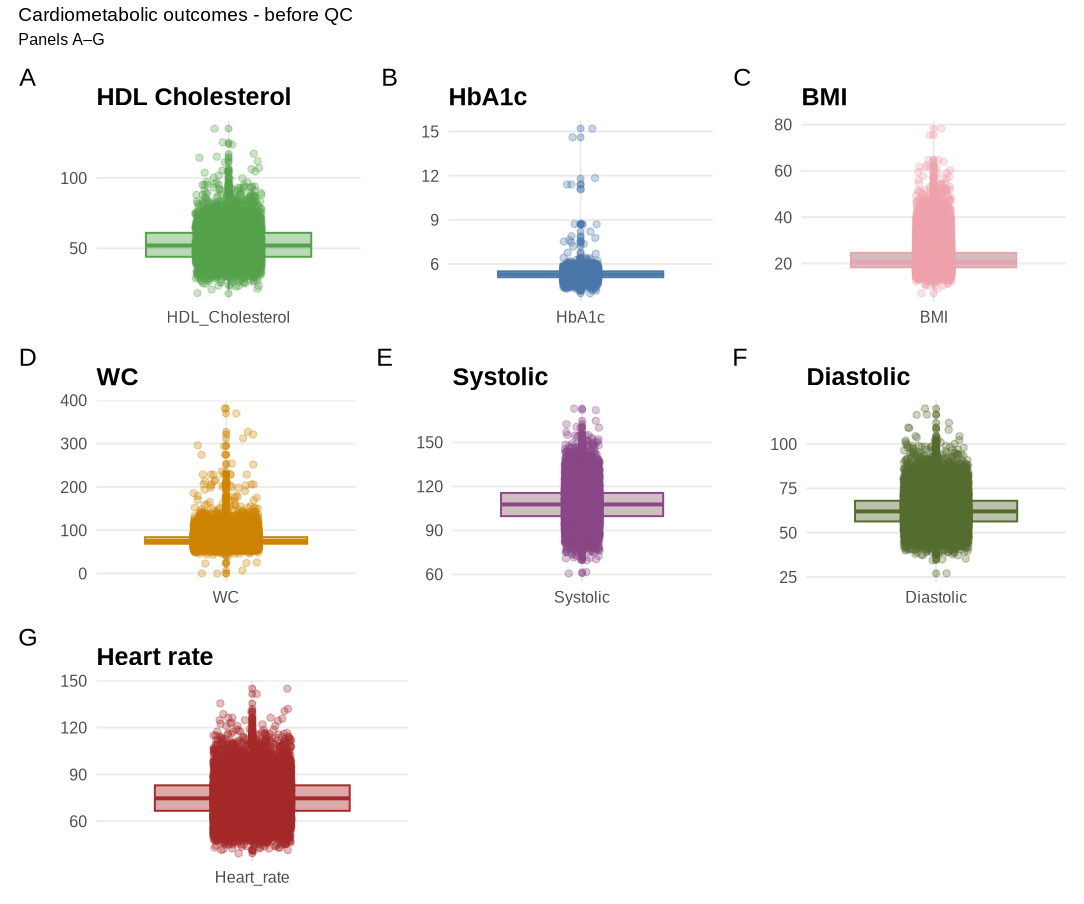

**SI Figure 8**. Showing cardiometabolic risk factors *before* quality assurance procedures.

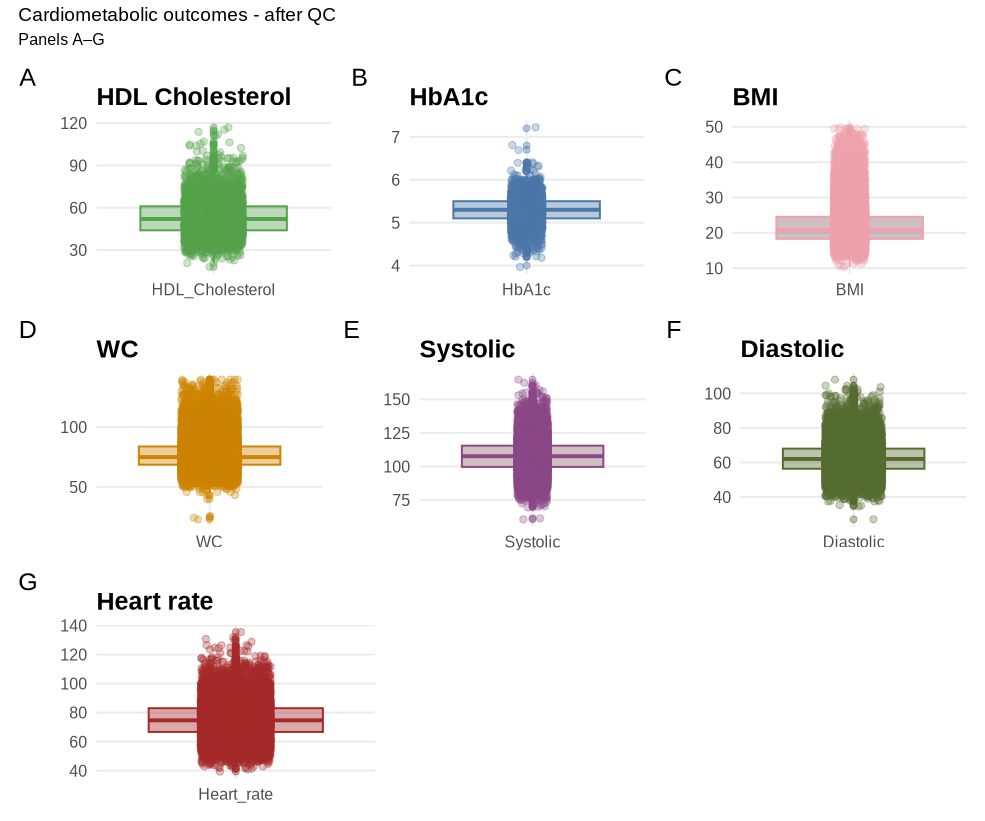

**SI Figure 9**. Showing cardiometabolic risk factors *after* quality assurance procedures.

| **SI Table 4.** **Bayes Factor (BF_12_).** Showing evidence ratio interpretations^14^. Values of 1 can be interpreted as no differential evidence, with the following values indicating weight of evidence in favour of the alternative hypothesis for model 2 (**M_2_**): 0.3-1 (anecdotal), 0.1-0.3 (moderate), 0.03-0.1 (strong), 0.01- 0.03 (very strong), <0.01 (extreme). Contrarily, the following values indicate weight of evidence in favour of the null hypothesis for model 1 (**M_1_**): 1-3 (anecdotal), 3-10 (moderate), 10-30 (strong), 30-100 (very strong), >100 (extreme). | | | |
| --- | --- | --- | --- |
| Bayes factor BF_12_ | | | Interpretation |
|  | > | 100 | Extreme evidence for M_1_ |
| 30 | - | 100 | Very strong evidence for M_1_ |
| 10 | - | 30 | Strong evidence for M_1_ |
| 3 | - | 10 | Moderate evidence for M_1_ |
| 1 | - | 3 | Anecdotal evidence for M_1_ |
|  | 1 |  | No evidence |
| 0.3 | - | 1 | Anecdotal evidence for M_2_ |
| 0.10 | - | 0.3 | Moderate evidence for M_2_ |
| 0.03 | - | 0.10 | Strong evidence for M_2_ |
| 0.01 | - | 0.03 | Very strong evidence for M_2_ |
|  | < | 0.01 | Extreme evidence for M_2_ |

**SI Table 5**. Results of the primary Bayesian multilevel models (CMR main effects and CMR×Timepoint interactions) for global brain MRI features.

| **Model** | **CMR feature** | **Estimate** | **lower95** | **upper95** | **P higher** | **P lower** | **BF₁₂** |
| --- | --- | --- | --- | --- | --- | --- | --- |
| CT | Systolic | 0 | -0.03 | 0.031 | 0.497 | 0.503 | 65.185 |
| CT | Diastolic | -0.022 | -0.049 | 0.007 | 0.067 | 0.933 | 22.958 |
| CT | Heart rate | -0.007 | -0.037 | 0.023 | 0.327 | 0.673 | 56.605 |
| CT | WC | -0.026 | -0.059 | 0.008 | 0.065 | 0.935 | 19.006 |
| CT | BMI | -0.049 | -0.084 | -0.015 | 0.002 | 0.998 | 0.996 |
| CT | Hba1c | -0.041 | -0.078 | -0.007 | 0.01 | 0.99 | 4.383 |
| CT | HDL Cholesterol | 0 | -0.033 | 0.034 | 0.509 | 0.491 | 58.906 |
| CT | TP:Systolic | 0.009 | -0.004 | 0.021 | 0.911 | 0.089 | 62.111 |
| CT | TP:Diastolic | -0.002 | -0.014 | 0.009 | 0.356 | 0.644 | 146.283 |
| CT | TP:Heart rate | -0.006 | -0.018 | 0.007 | 0.173 | 0.827 | 99.772 |
| CT | TP:WC | -0.007 | -0.021 | 0.007 | 0.154 | 0.846 | 81.116 |
| CT | TP:BMI | -0.015 | -0.029 | -0.001 | 0.017 | 0.983 | 15.639 |
| CT | TP:Hba1c | -0.011 | -0.026 | 0.003 | 0.073 | 0.927 | 43.687 |
| CT | TP:HDL Cholesterol | -0.006 | -0.02 | 0.009 | 0.227 | 0.773 | 104.877 |
| SA | Systolic | 0.012 | -0.003 | 0.026 | 0.946 | 0.054 | 38.431 |
| SA | Diastolic | -0.005 | -0.019 | 0.008 | 0.216 | 0.784 | 110.37 |
| SA | Heart rate | 0.004 | -0.011 | 0.019 | 0.703 | 0.297 | 111.335 |
| SA | WC | 0.034 | 0.015 | 0.054 | 1 | 0 | 0.265 |
| SA | BMI | 0.007 | -0.017 | 0.03 | 0.709 | 0.291 | 72.118 |
| SA | Hba1c | -0.006 | -0.028 | 0.016 | 0.304 | 0.696 | 80.246 |
| SA | HDL Cholesterol | -0.02 | -0.037 | -0.003 | 0.01 | 0.99 | 8.062 |
| SA | TP:Systolic | 0.004 | -0.002 | 0.01 | 0.928 | 0.072 | 115.666 |
| SA | TP:Diastolic | -0.003 | -0.009 | 0.003 | 0.143 | 0.857 | 188.355 |
| SA | TP:Heart rate | 0.004 | -0.003 | 0.01 | 0.885 | 0.115 | 158.842 |
| SA | TP:WC | 0.008 | 0 | 0.016 | 0.98 | 0.02 | 33.635 |
| SA | TP:BMI | -0.001 | -0.01 | 0.007 | 0.377 | 0.623 | 230.071 |
| SA | TP:Hba1c | -0.006 | -0.014 | 0.003 | 0.103 | 0.897 | 100.071 |
| SA | TP:HDL Cholesterol | -0.007 | -0.014 | 0.001 | 0.036 | 0.964 | 52.972 |
| FA | Systolic | 0.024 | -0.011 | 0.057 | 0.913 | 0.087 | 21.698 |
| FA | Diastolic | -0.003 | -0.036 | 0.028 | 0.431 | 0.569 | 59.184 |
| FA | Heart rate | -0.024 | -0.057 | 0.01 | 0.083 | 0.917 | 22.631 |
| FA | WC | -0.008 | -0.044 | 0.03 | 0.34 | 0.66 | 47.962 |
| FA | BMI | -0.017 | -0.053 | 0.021 | 0.18 | 0.82 | 33.843 |
| FA | Hba1c | -0.002 | -0.041 | 0.036 | 0.455 | 0.545 | 51.247 |
| FA | HDL Cholesterol | 0.008 | -0.029 | 0.046 | 0.656 | 0.344 | 47.773 |
| FA | TP:Systolic | 0.006 | -0.008 | 0.02 | 0.81 | 0.19 | 93.962 |
| FA | TP:Diastolic | -0.002 | -0.015 | 0.012 | 0.387 | 0.613 | 135.916 |
| FA | TP:Heart rate | -0.008 | -0.022 | 0.005 | 0.121 | 0.879 | 69.105 |
| FA | TP:WC | -0.001 | -0.017 | 0.014 | 0.437 | 0.564 | 122.868 |
| FA | TP:BMI | -0.005 | -0.021 | 0.009 | 0.247 | 0.753 | 101.96 |
| FA | TP:Hba1c | 0.002 | -0.014 | 0.019 | 0.578 | 0.422 | 118.483 |
| FA | TP:HDL Cholesterol | 0.002 | -0.014 | 0.018 | 0.585 | 0.415 | 121.134 |
| MD | Systolic | 0.001 | -0.03 | 0.033 | 0.535 | 0.465 | 61.499 |
| MD | Diastolic | 0.005 | -0.025 | 0.034 | 0.629 | 0.371 | 62.445 |
| MD | Heart rate | 0.066 | 0.035 | 0.099 | 1 | 0 | 0.045 |
| MD | WC | -0.021 | -0.056 | 0.013 | 0.115 | 0.885 | 27.722 |
| MD | BMI | -0.033 | -0.07 | 0.002 | 0.035 | 0.965 | 11.839 |
| MD | Hba1c | 0.003 | -0.033 | 0.039 | 0.568 | 0.432 | 50.857 |
| MD | HDL Cholesterol | -0.016 | -0.052 | 0.018 | 0.183 | 0.817 | 39.356 |
| MD | TP:Systolic | 0.006 | -0.007 | 0.02 | 0.801 | 0.199 | 107.976 |
| MD | TP:Diastolic | 0.005 | -0.008 | 0.018 | 0.765 | 0.235 | 114.745 |
| MD | TP:Heart rate | 0.027 | 0.014 | 0.04 | 1 | 0 | 0.068 |
| MD | TP:WC | -0.003 | -0.018 | 0.012 | 0.356 | 0.644 | 120.014 |
| MD | TP:BMI | -0.004 | -0.018 | 0.011 | 0.294 | 0.706 | 118.971 |
| MD | TP:Hba1c | 0.003 | -0.012 | 0.019 | 0.663 | 0.337 | 122.117 |
| MD | TP:HDL Cholesterol | -0.007 | -0.022 | 0.009 | 0.19 | 0.81 | 86.405 |

**Note**: All models have a normal prior of 1. Evidence: evidence ratio; CT: cortical thickness; SA: surface area; FA: fractional anisotropy; MD: mean diffusivity; Systolic BP: systolic blood pressure; Diastolic BP: diastolic blood pressure; BMI: body-mass index; WC: waist circumference; HDL Cholesterol: High-density-lipoprotein cholesterol; Hba1c: glycated haemoglobin; Pulse: resting heart rate; TP: timepoint.

**SI Table 6**. Results of the within-person **change-score** models (Δbrain ~ ΔCMR), showing standardised coefficients for all selected global brain MRI features.

| **Model** | **CMR feature** | **Estimate** | **lower95** | **upper95** | **P higher** | **P lower** | **BF₁₂** |
| --- | --- | --- | --- | --- | --- | --- | --- |
| CT | Systolic | 0 | -0.049 | 0.049 | 0.5 | 0.5 | 39.726 |
| CT | Diastolic | -0.032 | -0.077 | 0.014 | 0.083 | 0.917 | 16.857 |
| CT | Heart rate | -0.01 | -0.062 | 0.04 | 0.353 | 0.647 | 34.929 |
| CT | WC | 0.097 | 0.023 | 0.174 | 0.995 | 0.005 | 1.047 |
| CT | BMI | 0.066 | -0.039 | 0.163 | 0.898 | 0.102 | 8.892 |
| CT | Hba1c | 0.033 | -0.044 | 0.115 | 0.792 | 0.208 | 18.213 |
| CT | HDL Cholesterol | 0.005 | -0.055 | 0.063 | 0.565 | 0.435 | 32.636 |
| SA | Systolic | 0.009 | -0.007 | 0.024 | 0.864 | 0.136 | 71.169 |
| SA | Diastolic | 0 | -0.014 | 0.015 | 0.504 | 0.496 | 134.259 |
| SA | Heart rate | 0.003 | -0.014 | 0.018 | 0.627 | 0.373 | 117.09 |
| SA | WC | 0.03 | 0.007 | 0.054 | 0.994 | 0.006 | 4.33 |
| SA | BMI | 0.028 | -0.006 | 0.059 | 0.956 | 0.044 | 14.544 |
| SA | Hba1c | 0.003 | -0.021 | 0.028 | 0.588 | 0.412 | 76.359 |
| SA | HDL Cholesterol | -0.012 | -0.03 | 0.005 | 0.087 | 0.913 | 43.522 |
| FA | Systolic | 0.024 | -0.037 | 0.084 | 0.78 | 0.22 | 24.584 |
| FA | Diastolic | -0.016 | -0.071 | 0.039 | 0.292 | 0.708 | 31.307 |
| FA | Heart rate | 0.034 | -0.03 | 0.096 | 0.854 | 0.146 | 19.027 |
| FA | WC | -0.053 | -0.144 | 0.04 | 0.133 | 0.867 | 11.214 |
| FA | BMI | 0.006 | -0.124 | 0.13 | 0.536 | 0.464 | 14.896 |
| FA | Hba1c | 0.042 | -0.053 | 0.135 | 0.81 | 0.19 | 13.925 |
| FA | HDL Cholesterol | 0.051 | -0.022 | 0.127 | 0.912 | 0.088 | 10.649 |
| MD | Systolic | -0.006 | -0.062 | 0.048 | 0.41 | 0.59 | 34.399 |
| MD | Diastolic | 0.036 | -0.015 | 0.087 | 0.914 | 0.086 | 15.168 |
| MD | Heart rate | 0.053 | -0.007 | 0.108 | 0.964 | 0.036 | 7.373 |
| MD | WC | 0.071 | -0.014 | 0.158 | 0.947 | 0.053 | 6.133 |
| MD | BMI | 0.028 | -0.086 | 0.144 | 0.685 | 0.315 | 15.715 |
| MD | Hba1c | -0.013 | -0.107 | 0.074 | 0.39 | 0.61 | 20.779 |
| MD | HDL Cholesterol | -0.08 | -0.147 | -0.013 | 0.01 | 0.99 | 2.23 |

**SI Table 7**. Results of the lagged prospective models (brain _𝑡 + 1_ ~ CMR _𝑡_ + brain _𝑡_), showing standardised coefficients for all selected global brain MRI features.

| **Model** | **CMR feature** | **Estimate** | **lower95** | **upper95** | **P higher** | **P lower** | **BF₁₂** |
| --- | --- | --- | --- | --- | --- | --- | --- |
| CT | Systolic | 0.005 | -0.046 | 0.055 | 0.581 | 0.419 | 38.158 |
| CT | Diastolic | 0.023 | -0.031 | 0.074 | 0.807 | 0.192 | 25.052 |
| CT | Heart rate | 0.042 | -0.015 | 0.097 | 0.932 | 0.068 | 12.17 |
| CT | WC | -0.023 | -0.081 | 0.035 | 0.224 | 0.776 | 24.105 |
| CT | BMI | -0.038 | -0.101 | 0.025 | 0.117 | 0.883 | 15.274 |
| CT | Hba1c | -0.045 | -0.101 | 0.013 | 0.066 | 0.934 | 11.601 |
| CT | HDL Cholesterol | -0.021 | -0.07 | 0.025 | 0.199 | 0.801 | 28.977 |
| SA | Systolic | -0.015 | -0.03 | 0.001 | 0.029 | 0.971 | 19.979 |
| SA | Diastolic | -0.008 | -0.024 | 0.009 | 0.177 | 0.823 | 77.323 |
| SA | Heart rate | 0.011 | -0.006 | 0.028 | 0.895 | 0.105 | 51.673 |
| SA | WC | -0.017 | -0.036 | 0 | 0.03 | 0.97 | 16.965 |
| SA | BMI | -0.016 | -0.036 | 0.003 | 0.05 | 0.95 | 26.152 |
| SA | Hba1c | -0.002 | -0.02 | 0.016 | 0.394 | 0.606 | 108.753 |
| SA | HDL Cholesterol | 0.016 | 0.001 | 0.032 | 0.979 | 0.021 | 14.999 |
| FA | Systolic | -0.035 | -0.096 | 0.025 | 0.124 | 0.876 | 16.537 |
| FA | Diastolic | -0.035 | -0.097 | 0.028 | 0.14 | 0.86 | 17.55 |
| FA | Heart rate | -0.041 | -0.109 | 0.026 | 0.117 | 0.882 | 14.859 |
| FA | WC | 0.043 | -0.027 | 0.111 | 0.885 | 0.115 | 13.964 |
| FA | BMI | 0.054 | -0.02 | 0.128 | 0.924 | 0.076 | 9.657 |
| FA | Hba1c | 0.026 | -0.041 | 0.091 | 0.778 | 0.222 | 21.63 |
| FA | HDL Cholesterol | -0.035 | -0.094 | 0.024 | 0.12 | 0.88 | 16.725 |
| MD | Systolic | 0.074 | 0.018 | 0.126 | 0.996 | 0.004 | 1.244 |
| MD | Diastolic | 0.055 | -0.001 | 0.112 | 0.972 | 0.028 | 5.853 |
| MD | Heart rate | 0.013 | -0.045 | 0.075 | 0.668 | 0.332 | 30.081 |
| MD | WC | -0.013 | -0.073 | 0.051 | 0.346 | 0.655 | 28.537 |
| MD | BMI | -0.005 | -0.072 | 0.06 | 0.438 | 0.562 | 28.911 |
| MD | Hba1c | 0.051 | -0.007 | 0.113 | 0.953 | 0.047 | 7.959 |
| MD | HDL Cholesterol | 0.048 | -0.006 | 0.103 | 0.958 | 0.042 | 8.345 |

**SI Table 8.** Results of the **PDS-adjusted** Bayesian multilevel models (PDS in place of age), including CMR main effects and CMR×Timepoint interactions for selected global brain MRI features.

| **Model** | **CMR feature** | **Estimate** | **lower95** | **upper95** | **P higher** | **P lower** | **BF₁₂** |
| --- | --- | --- | --- | --- | --- | --- | --- |
| CT | Systolic | 0.002 | -0.028 | 0.033 | 0.557 | 0.443 | 61.727 |
| CT | Diastolic | -0.02 | -0.048 | 0.01 | 0.094 | 0.906 | 28.355 |
| CT | Heart rate | 0.001 | -0.029 | 0.033 | 0.541 | 0.459 | 61.612 |
| CT | WC | -0.018 | -0.052 | 0.018 | 0.154 | 0.846 | 34.141 |
| CT | BMI | -0.04 | -0.076 | -0.004 | 0.015 | 0.985 | 5.5 |
| CT | Hba1c | -0.038 | -0.075 | -0.003 | 0.019 | 0.981 | 6.284 |
| CT | HDL Cholesterol | 0.003 | -0.032 | 0.036 | 0.565 | 0.435 | 55.344 |
| CT | TP:Systolic | 0.009 | -0.004 | 0.021 | 0.911 | 0.089 | 65.417 |
| CT | TP:Diastolic | -0.002 | -0.014 | 0.01 | 0.364 | 0.636 | 150.904 |
| CT | TP:Heart rate | -0.003 | -0.015 | 0.01 | 0.34 | 0.66 | 142.738 |
| CT | TP:WC | -0.006 | -0.02 | 0.009 | 0.2 | 0.8 | 96.259 |
| CT | TP:BMI | -0.014 | -0.028 | 0 | 0.027 | 0.973 | 22.718 |
| CT | TP:Hba1c | -0.011 | -0.026 | 0.005 | 0.082 | 0.918 | 49.87 |
| CT | TP:HDL Cholesterol | -0.004 | -0.02 | 0.01 | 0.304 | 0.696 | 110.16 |
| SA | Systolic | 0.009 | -0.006 | 0.024 | 0.879 | 0.121 | 64.684 |
| SA | Diastolic | -0.007 | -0.02 | 0.007 | 0.162 | 0.838 | 88.27 |
| SA | Heart rate | 0.008 | -0.007 | 0.024 | 0.84 | 0.16 | 78.415 |
| SA | WC | 0.032 | 0.012 | 0.052 | 0.999 | 0.001 | 0.65 |
| SA | BMI | 0.004 | -0.02 | 0.029 | 0.625 | 0.375 | 74.701 |
| SA | Hba1c | -0.004 | -0.025 | 0.017 | 0.372 | 0.628 | 93.048 |
| SA | HDL Cholesterol | -0.017 | -0.034 | 0 | 0.025 | 0.975 | 17.488 |
| SA | TP:Systolic | 0.004 | -0.002 | 0.01 | 0.901 | 0.099 | 148.15 |
| SA | TP:Diastolic | -0.003 | -0.009 | 0.003 | 0.144 | 0.856 | 201.959 |
| SA | TP:Heart rate | 0.005 | -0.001 | 0.011 | 0.941 | 0.059 | 101.69 |
| SA | TP:WC | 0.008 | 0 | 0.016 | 0.976 | 0.024 | 35.224 |
| SA | TP:BMI | -0.002 | -0.01 | 0.007 | 0.359 | 0.641 | 224.177 |
| SA | TP:Hba1c | -0.005 | -0.013 | 0.004 | 0.149 | 0.851 | 136.539 |
| SA | TP:HDL Cholesterol | -0.006 | -0.013 | 0.001 | 0.058 | 0.942 | 78.693 |
| FA | Systolic | 0.026 | -0.008 | 0.062 | 0.932 | 0.068 | 19.08 |
| FA | Diastolic | -0.002 | -0.034 | 0.031 | 0.453 | 0.547 | 59.654 |
| FA | Heart rate | -0.031 | -0.066 | 0.003 | 0.039 | 0.961 | 11.729 |
| FA | WC | -0.009 | -0.046 | 0.029 | 0.327 | 0.673 | 46.592 |
| FA | BMI | -0.021 | -0.059 | 0.018 | 0.145 | 0.855 | 30.282 |
| FA | Hba1c | -0.006 | -0.046 | 0.032 | 0.376 | 0.624 | 47.22 |
| FA | HDL Cholesterol | 0.008 | -0.03 | 0.045 | 0.662 | 0.338 | 48.243 |
| FA | TP:Systolic | 0.007 | -0.007 | 0.021 | 0.853 | 0.147 | 80.417 |
| FA | TP:Diastolic | -0.002 | -0.015 | 0.012 | 0.412 | 0.588 | 148.139 |
| FA | TP:Heart rate | -0.011 | -0.025 | 0.003 | 0.058 | 0.942 | 39.425 |
| FA | TP:WC | 0 | -0.016 | 0.015 | 0.477 | 0.523 | 124.895 |
| FA | TP:BMI | -0.005 | -0.021 | 0.01 | 0.26 | 0.74 | 101.064 |
| FA | TP:Hba1c | 0 | -0.016 | 0.016 | 0.508 | 0.492 | 120.832 |
| FA | TP:HDL Cholesterol | 0.002 | -0.015 | 0.019 | 0.572 | 0.428 | 115.266 |
| MD | Systolic | 0.003 | -0.031 | 0.034 | 0.565 | 0.435 | 61.642 |
| MD | Diastolic | 0.005 | -0.025 | 0.036 | 0.635 | 0.365 | 59.289 |
| MD | Heart rate | 0.071 | 0.039 | 0.103 | 1 | 0 | 0 |
| MD | WC | -0.016 | -0.051 | 0.021 | 0.197 | 0.803 | 36.617 |
| MD | BMI | -0.026 | -0.063 | 0.011 | 0.084 | 0.916 | 21.383 |
| MD | Hba1c | 0.005 | -0.032 | 0.043 | 0.609 | 0.391 | 49.142 |
| MD | HDL Cholesterol | -0.019 | -0.053 | 0.018 | 0.141 | 0.859 | 31.33 |
| MD | TP:Systolic | 0.005 | -0.008 | 0.019 | 0.773 | 0.227 | 112.302 |
| MD | TP:Diastolic | 0.004 | -0.008 | 0.017 | 0.746 | 0.254 | 123.591 |
| MD | TP:Heart rate | 0.029 | 0.017 | 0.043 | 1 | 0 | 0.005 |
| MD | TP:WC | -0.003 | -0.018 | 0.011 | 0.365 | 0.635 | 126.042 |
| MD | TP:BMI | -0.004 | -0.019 | 0.01 | 0.313 | 0.687 | 119.61 |
| MD | TP:Hba1c | 0.003 | -0.013 | 0.019 | 0.638 | 0.362 | 119.549 |
| MD | TP:HDL Cholesterol | -0.008 | -0.024 | 0.008 | 0.175 | 0.825 | 82.337 |

**Note**: Replacing age with pubertal stage (PDS) left the overall pattern unchanged. For cortical thickness, most cardiometabolic predictors favoured the null; the BMI–CT association was null (β = −0.040, 95% HDI [−0.076, −0.004]; BF₁₂ = 5.500, moderate evidence for M1), and other CT effects and interactions were similarly null including the BMI×TP interaction (β = −0.014, [−0.028, 0.000]; BF₁₂ = 22.718, strong evidence for M1). Surface area showed a positive association with waist circumference (β = 0.032, [0.012, 0.052]; BF₁₂ = 0.650, anecdotal evidence for M2), while all other surface area and fractional anisotropy effects showed consistent null results across main effects and TP interactions (strong–extreme evidence for M1). For mean diffusivity, resting heart rate remained robustly positive (β = 0.071, [0.039, 0.103]; BF₁₂ < 0.001, extreme evidence for M2) and strengthened across time (Heart rate × TP β = 0.029 per wave, [0.017, 0.043]; BF₁₂ = 0.005, very strong evidence for M2); other MD predictors favoured the null.

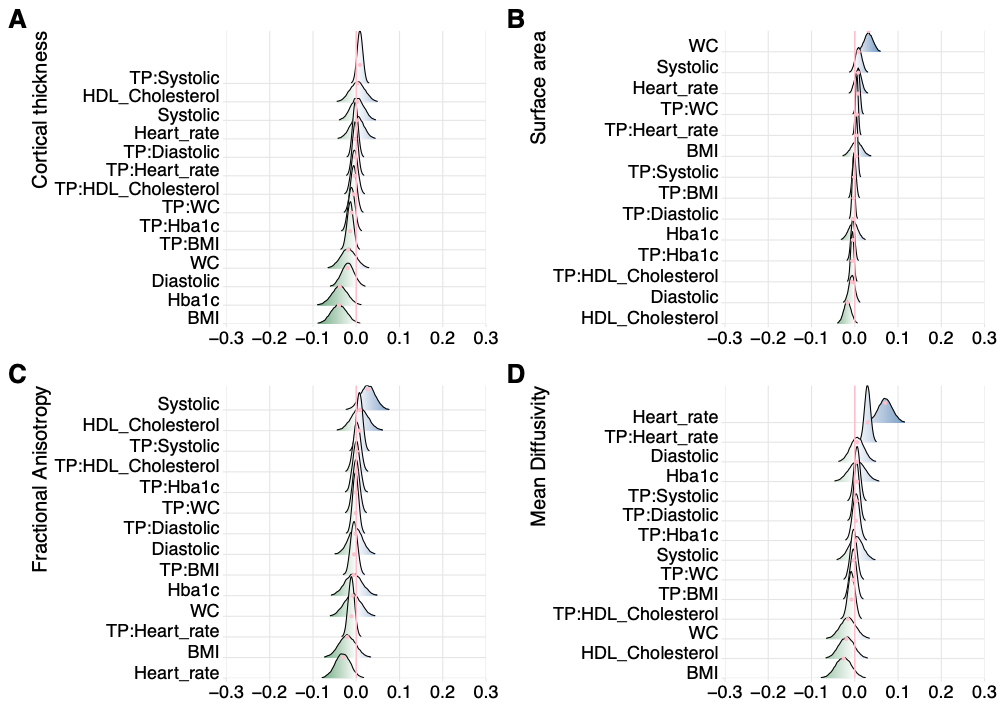

**SI Figure 10**. **PDS-adjusted associations between CMRs and global brain structure.** Posterior distributions of standardised coefficients from the Bayesian multilevel model with pubertal development (PDS) in place of age (sex and SES retained). Panels show effects for (A) cortical thickness, (B) surface area, (C) fractional anisotropy, and (D) mean diffusivity, including CMR main effects and CMR × Timepoint interactions. Colour indicates direction (blue = positive; green = negative); distribution width reflects uncertainty. Systolic BP: systolic blood pressure; Diastolic BP: diastolic blood pressure; Heart rate: heart rate; HDL Cholesterol: high-density lipoprotein cholesterol; Haemoglobin A1c: HbA1c; BMI: body-mass index; WC: waist circumference; TP: timepoint.

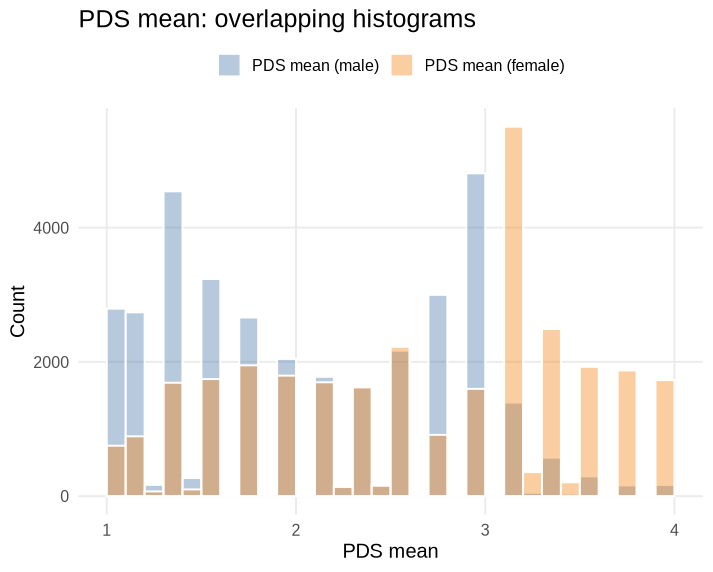

**SI Figure 11.** Showing PDS sum scores per sex. The PDS includes five-items, each rated on a 4-point scale (1 = no development; 2 = development has barely begun; 3 = development is definitely underway; and 4 = development is complete; except menstruation, which is coded 1 = has not begun, 4 = has begun). Thus, higher scores reflect more advanced pubertal development.
